# Habitual and supplemented prebiotic diets and their links to inflammatory serum markers and hypothalamic microstructure in young, overweight adults: a pre-registered study

**DOI:** 10.1101/2023.11.01.23297892

**Authors:** Emmy Töws, Evelyn Medawar, Ronja Thieleking, Frauke Beyer, Michael Stumvoll, Arno Villringer, A. Veronica Witte

## Abstract

**Background:** Prebiotic dietary fiber and related metabolites have been suggested to attenuate low-grade systemic and central inflammation through improving gut-brain axis signaling. We here aimed to test whether habitual or short-term high-dose fiber intake is linked to inflammatory markers in blood and to indicators of central hypothalamic inflammation.

**Methods:** In total, 59 adults (19 women, aged 28.3 years ± 6.6 SD, mean body mass index, BMI, 27.3 ± 1.5 SD) were included into analyses. Participants completed a food frequency questionnaire, underwent diffusion-weighted magnetic resonance imaging (MRI) at 3 Tesla for provision of mean diffusivity (MD) as a marker of brain tissue inflammation and donated fasting blood. Measurements took place at up to 4 timepoints, i.e. before and after 14 days of supplementary fiber and placebo intake, respectively. High-sensitive C-reactive protein (CRP), tumor-necrosis factor alpha (TNF-α) and interleukin-6 (IL6) were assessed in serum. The study was preregistered at https://osf.io/uzbav.

**Results:** Habitual and interventional high-fiber diet was not significantly associated with neither inflammatory markers (|ß_intervention_|> 0.1, p > 0.32) nor with hypothalamic MD (|ß_intervention_| = 1.8, p = 0.07) according to linear mixed effects modeling. Male sex and higher body fat mass related to higher CRP. Further, higher BMI was borderline related to lower hypothalamic MD.

**Conclusions:** In this sample of overweight adults, dietary fiber intake was not related to inflammatory blood markers or hypothalamic microstructure. Instead, sex and body composition were of higher importance for prediction of interindividual differences in markers of (neuro)inflammation.

**Significance Statement:** Prebiotic dietary fiber has been discussed to lower systemic and central inflammation. While previous studies investigated the effects of fiber on inflammatory blood markers, the knowledge of the effect of fiber on neuroinflammation is limited. Thus, in this pre-registered randomized controlled trial analysis we examined the relationship between dietary fiber intake and inflammatory markers in blood and hypothalamus. 3T MRI and blood markers were assessed before and after high-fiber intake and placebo in 59 adults. In our overweight study sample of 19-42 years old adults, fiber intake had no significant impact on inflammatory markers. The current null findings can inform future nutrition neuroimaging trials and add to the discussion about how diet may affect brain structure and function.

## Introduction

High-fat diet and fat accumulation can trigger low-grade systemic inflammation, leading to maladaptive changes in food intake-related brain areas such as the hypothalamus (Thaler and Schwartz 2010, Sewaybricker et al., 2023). Diets high in saturated fatty acids may activate inflammatory pathways (Rocha et al. 2016) and modulate intestinal microbiota. Thereby, they increase intestinal permeability and induce systemic inflammation (Cani et al. 2008; Deopurkar et al. 2010; Lassenius et al. 2011). Consequently, inflammatory serum factors which cross the blood-brain barrier can provoke dysfunctional changes in brain areas such as the hypothalamus (Van Dyken and Lacoste 2018).

In contrast to high-fat diets, high-fiber diets have been discussed to exert anti-inflammatory effects in gut and circulation (Dalile et al. 2019; Medawar et al. 2019). Dietary fibers are converted into short-chain fatty acids (SCFAs) through bacterial fermentation in the colon. SCFAs alleviate inflammatory processes at their production site in the colon and systemically after entering blood circulation (Morrison and Preston 2016).

In the gut, SCFA contribute to a decreased permeability of the intestinal membrane by facilitating tight-junction-assembly. Thus, bacteria expressing proinflammatory lipopolysaccharides on their surface are prevented from entering extraintestinal circulation (Luying Peng, Zhong-Rong Li, Robert S. Green, Ian R. Holzman 2009). Hence, extraintestinal inflammation initiated by bacterial components can be contained. Additionally, SCFAs have been suggested to act via the immune pathway influencing systemic and neuroinflammation (Dalile et al. 2019).

Moreover, dietary fiber or its derivative SCFAs promote the secretion of gut-derived anorexigenic hormones, such as glucagon-like peptide-1 (GLP-1) and peptide YY (PYY). These hormones induce satiety via the suppression of appetite-stimulating hypothalamic neuropeptide Y (NPY) neurons and the activation of appetite-suppressing pro-opiomelanocortin (POMC) neurons in the hypothalamus (De Silva and Bloom 2012). Thereby, SCFAs may contribute to higher satiety. Higher satiety in turn may lead to less high-fat food consumption (Byrne et al. 2015) resulting in lower inflammatory parameters. An alleviation of inflammatory processes may potentially link to higher functional hypothalamic integrity and therefore a more sensitive regulation of appetite.

Human, cross-sectional epidemiological studies report that higher habitual high-fiber diet links to lower levels of peripheral inflammatory markers (Ma et al. 2008; Mazidi et al. 2018; Wannamethee et al. 2009). Interventional studies which examine a potential association between fiber intake and inflammatory processes are however scarce and limited in sample size. One randomized clinical interventional study investigated 35 (18 obese and 17 lean) individuals following either a “Dietary Approaches to Stop Hypertension” diet (DASH, a diet rich in fiber and low in dairy and saturated fat) or a high-fiber supplementation diet (30g/day of psyllium) for 3 weeks respectively in a cross-over design (King et al. 2007). Although both diets reduced C-reactive protein (CRP) levels, the fiber supplement showed slightly stronger effects than the DASH diet. Notably, only lean participants showed an amelioration of inflammatory markers. This clinical interventional study indicates that fiber may causally lower inflammatory markers, especially in lean individuals.

In sum, previous studies imply that dietary factors relate to (neuro-)inflammation in distinct ways, and some suggest prebiotic dietary fiber as anti-inflammatory agent. Yet, it remains unclear if a high-fiber diet relates to lower systemic and hypothalamic inflammation in non-lean individuals. Therefore, this randomized controlled study in a homogenous, well-characterized cohort of overweight adults aims to investigate whether dietary fiber exerts beneficial effects on markers of systemic inflammation and hypothalamic microstructure.

According to pre-registration (https://osf.io/uzbav), we hypothesized that higher habitual dietary fiber intake, measured using self-report of dietary habits over the course of seven days, correlates with (1) lower levels of the inflammatory markers interleukin-6 (IL-6), CRP, and tumor-necrosis factor alpha (TNF-a) in blood, and (2) with lower microstructural coherence in the hypothalamus measured using mean diffusivity (MD) derived from diffusion-weighted magnetic resonance imaging (MRI). In addition, we hypothesized that a two-week prebiotic fiber intervention (30 g inulin/d) would lead to improvements in blood (3) and brain markers (4).

## Material and Methods

### Ethics Approval and Recruitment

This study is part of a within-subject cross-over randomized controlled trial (RCT) investigating the effects of a prebiotic intervention on the gut-brain axis (Medawar et al., Gut, in press). The institutional Ethics Board of the Medical Faculty of the University of Leipzig, Germany, raised no concerns regarding the study protocol (228/18-ek) and all participants provided written informed consent. Recruitment took place via the institute’s database and advertisements. Remuneration was 9-10€/h and an additional bonus payment of 30€ for completing the study. The RCT was registered at ClinicalTrials.gov (#NCT03829189) and this analysis was pre-registered at https://osf.io/uzbav.

### Study Population

Out of 106 screened individuals we included a sample of 59 overweight adults (19 females, 40 males), aged 19-42 years (28 years ± 6.2 SD, BMI range 25-30 kg/m^2^, mean 27.3 kg/m^2^ ± 1.4 SD), for a flowchart, see **Extended Figure 1-1**. All participants assigned to either being female or male (alternative options: diverse, preferring not to report). Due to anatomical differences between females and males, we referred to female and male ‘sex’ considering differences in anthropometrics or brain morphology. Female and male ‘gender’ was used in all other occasions.

### Inclusion/Exclusion criteria

Inclusion criteria for this study were an age range of 18-45 years, a BMI of 25-30 kg/m^2^ upon first baseline assessment, no MRI contraindications, an omnivorous, non-restrictive diet, and no food allergies. Further any type of diet or antibiotic treatment in the last 3 months led to exclusion. Additionally, female participants had to regularly use oral or alternative contraceptives to minimize hormonal variations induced by the menstrual cycle. Pregnant and lactating women were not allowed to take part in the study. Participants were excluded if they suffered from a diagnosed neurological, psychiatric, or metabolic disorder. Diseases of the gastrointestinal tract, cardiovascular system, lung, liver, or kidneys led to exclusion as well as the intake of medication acting on the central nervous system. Daily alcohol intake had to be at a maximum of 50 grams. Limits for cigarette and coffee consumption were set to 10 cigarettes and 6 cups of coffee per day. Participants were dropouts when the supplement intake was missed out for more than 48 hours or if more than half of the 26 portions were missed out.

### Study Design

Participants underwent up to four assessments over the course of ∼6-10 weeks and in-between two assessments, they supplemented their diet with high-dosed prebiotic fiber (2 sachets of 15 g inulin/d), and with placebo (2 sachets isocaloric maltodextrin), respectively for 14 days (**Figure 1**). Specifically, daily intake of 30 g inulin contained 63 kcal and 26.7 g fiber (Orafti® Beneo Synergy1, BENEO GmbH, Mannheim, Germany) and placebo intake consisted of 16 g maltodextrin (63 kcal, 0 g fiber). Randomized allocation to study arm 1 or 2 determined the order of supplement intake. A wash-out period of at least 14 days was set between each of the interventions to avoid carry-over effects from the first intervention on the second.

**Figure 1:**
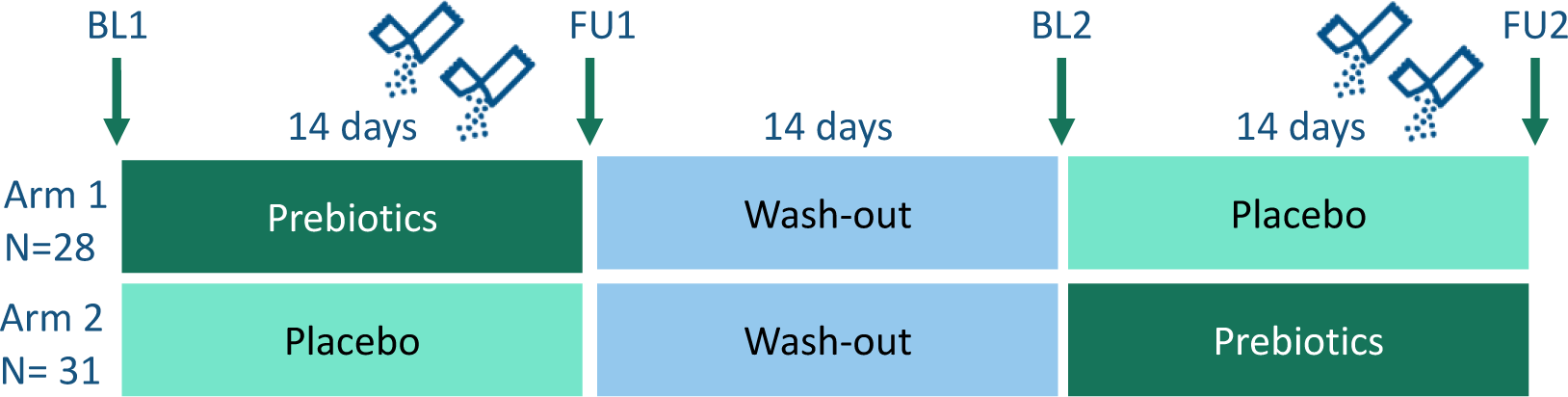
Study design. Each participant underwent up to four assessments: Baseline 1 (BL1, before first intervention), Follow-up 1 (FU1, after first intervention), Baseline 2 (before second intervention), Follow-up 2 (after second intervention). In-between two assessments participants supplemented their diet either with “prebiotics” (30g of inulin) or “placebo” (isocaloric maltodextrin). Participants were randomly allocated to study arm 1 or 2 which determined the order of supplement intake. A washout period of at least 14 days was set between interventions.

During the first three days of the intervention period the participants were asked to take one portion (15 g) of the supplement daily. Starting on day 4 and continuing through day 14, the amount of intake consisted of two portions daily (30 g). On day 15 (day of measurement) one sachet was added to a standardized breakfast shake after overnight fast and blood draw, so that one intervention period included 26 portions. We recommended to take the supplement before 5 p.m. to achieve a proper digestion before sleep. Fasting blood samples, anthropometrics, dietary habits and MRI were acquired on all four assessment days.

### Inflammatory markers

Participants were asked to fast from the evening prior to the blood drawing (mean 12.5 h ± 2.2 SD). The fasting blood samples were collected by trained staff at the same time point for each of the four measurements (using safety-multifly needles (21G, 200mm)). Blood samples were centrifuged at 3,500 revolutions per minute at 7°C for 6 minutes and the serum was aliquoted within one hour of obtainment. Processed aliquots were stored in a −80°C freezer within one hour of collection until the study was completed to analyze all samples in one batch. Analysis was conducted by Synevo Studien Service Labor GmbH c/o IMD Institut für Medizinische Diagnostik Berlin-Potsdam GbR, Berlin, Germany. We measured Il-6, CRP, and TNF-α.

### Anthropometric data

Body mass index (BMI) was measured as body weight (kg) divided by squared body height (m^2^). Participants were weighed in light clothes and without shoes in a fasted state on the same weight scale (100 g resolution, Seca GmbH, Germany) and their height was measured while standing against the wall with a fixed measuring scale (0.5 cm resolution, Seca GmbH, Germany). Percent body fat mass was measured using bioimpedance analysis with BIACORPUS RX 4004M (Medi Cal Healthcare GmbH, Karlsruhe, Germany) and two electrodes each at both hands and feet. Body fat mass was sex-standardized using z-transformation before analysis.

### Habitual dietary fiber intake

For estimation of the amount of habitual dietary fiber intake, participants were asked to complete the validated German food frequency questionnaire (DEGS1 FFQ) (Haftenberger et al. 2010) at each assessment. An in-house scoring tool was used to estimate the consumption of single food items and resulting daily nutrient intake based on self-report of frequency and quantity within seven days (Thieleking et al. 2023). We measured the amount of fiber intake using two different units: fiber in grams per day (absolute intake) and fiber per 1000kcal per day (relative intake) to adjust for overall caloric intake.

### MRI acquisition

MRI was performed on a 3T Siemens Prismafit scanner with a 32-channel head coil. MRI was acquired using a T1-weighted MPRAGE sequence using the ADNI protocol with the following parameters: TR = 2300ms; TE = 2.98ms; flip angle = 9°; FOV: (256 mm)²; voxel size: (1.0mm)³; 176 slices. Diffusion-weighted MRI (dwMRI) was acquired using the following parameters: TR = 5200ms; TE = 75ms; flip angle = 90°; FOV: (220 mm)²; voxel size: (1.7mm)³; 88 slices; max. b=1000 s/mm² in 60 diffusion directions; partial Fourier=7/8; GRAPPA-factor = 2; interpolation = OFF. Ap/pa-encoded b0-images were acquired for distortion correction.

### MRI data preprocessing

Anatomical images were automatically processed with the FreeSurfer v6.0.0p1 longitudinal stream, total intracranial volume per person and per time point was extracted based on the unbiased within-subject template space and image (Reuter et al. 2012). Several processing steps, such as skull stripping, Talairach transforms, atlas registration as well as spherical surface maps and parcellations were then initialized with common information from the within-subject template, to increase reliability and statistical power.

DwMRI preprocessing was performed with standard pipelines, including denoising (MRtrix v3.0; (Veraart, Fieremans, and Novikov 2016) of the raw data, removal of Gibbs-ringing artifact from all b0-images using the local subvoxel-shift method (Kellner et al. 2016) and outlier replacement using the EDDY tool (Andersson et al. 2016; Andersson and Sotiropoulos 2016) in FSL 6.0.1. (Smith et al. 2004). Subsequently, data was corrected for susceptibility distortions using topup (FSL) (Andersson, Skare, and Ashburner 2003). Brain masks of the unwarped b0-images were created using BET (Smith 2002) from FSL to correct for head motion and eddy currents using the EDDY tool (FSL; Bastiani et al. 2019). We applied tensor model fitting using DTIFit (FSL) to generate mean diffusivity (MD).

### Quality control of preprocessed DWI data

Using EDDY QC tools (FSL 6.0.1), quality control on person-wise and group-wise level have been performed with EDDY QUAD and EDDY SQUAD v1.0.2, respectively (Bastiani et al. 2019). The group-wise QC metrics (motion parameters, eddy currents, signal-to-noise ratio (SNR) and contrast-to-noise ratio (CNR)) have been compared to standard values (see **Table 1, Extended Fig. 2-1, 2-2**). Based on this assessment, we did not exclude any participants from analysis.

**Table 1:**
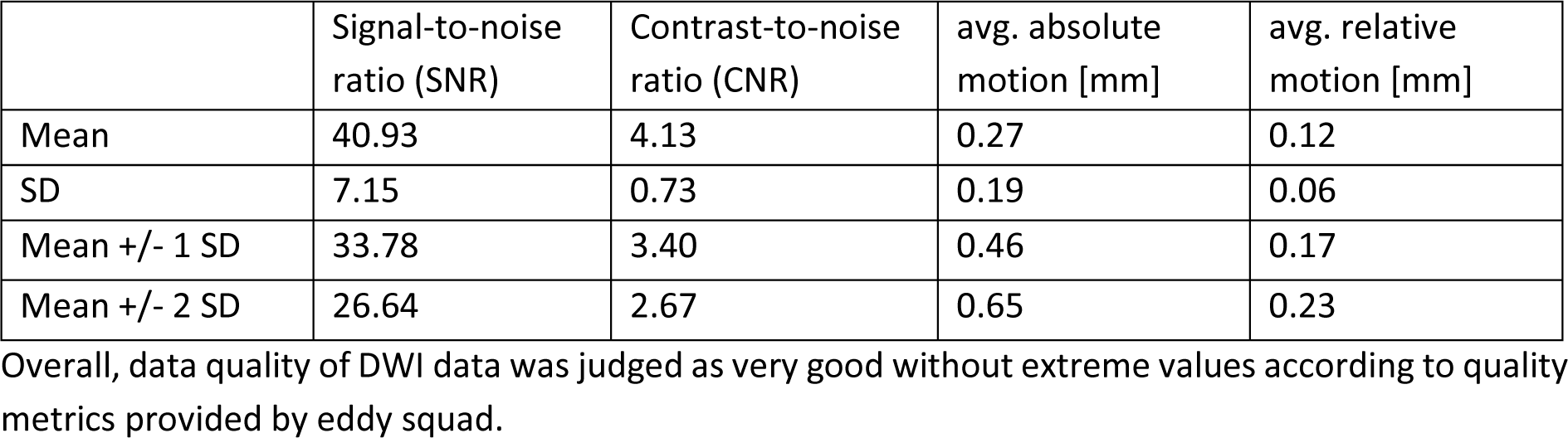
Diffusion-weighted imaging (DWI) Quality Control. Group-wise quality metrics provided by eddy squad for DWI data.

### Hypothalamus segmentation

Deviant to the pre-registration, we used automated segmentation of the bilateral hypothalamus using a deep learning algorithm (Billot et al. 2020) implemented in python scripts (v3.6) due to faster and more reliable results. Briefly, four hypothalamic regions were segmented at each hemisphere (Figure 2). Visual checks for correct segmentation on the structural image were done for all subjects. Volumes for total bilateral hypothalamus were extracted for each individual and for each of the up to four datapoints. Subnuclei were disregarded for statistical inference analysis to reduce Type II errors.

**Fig. 2:**
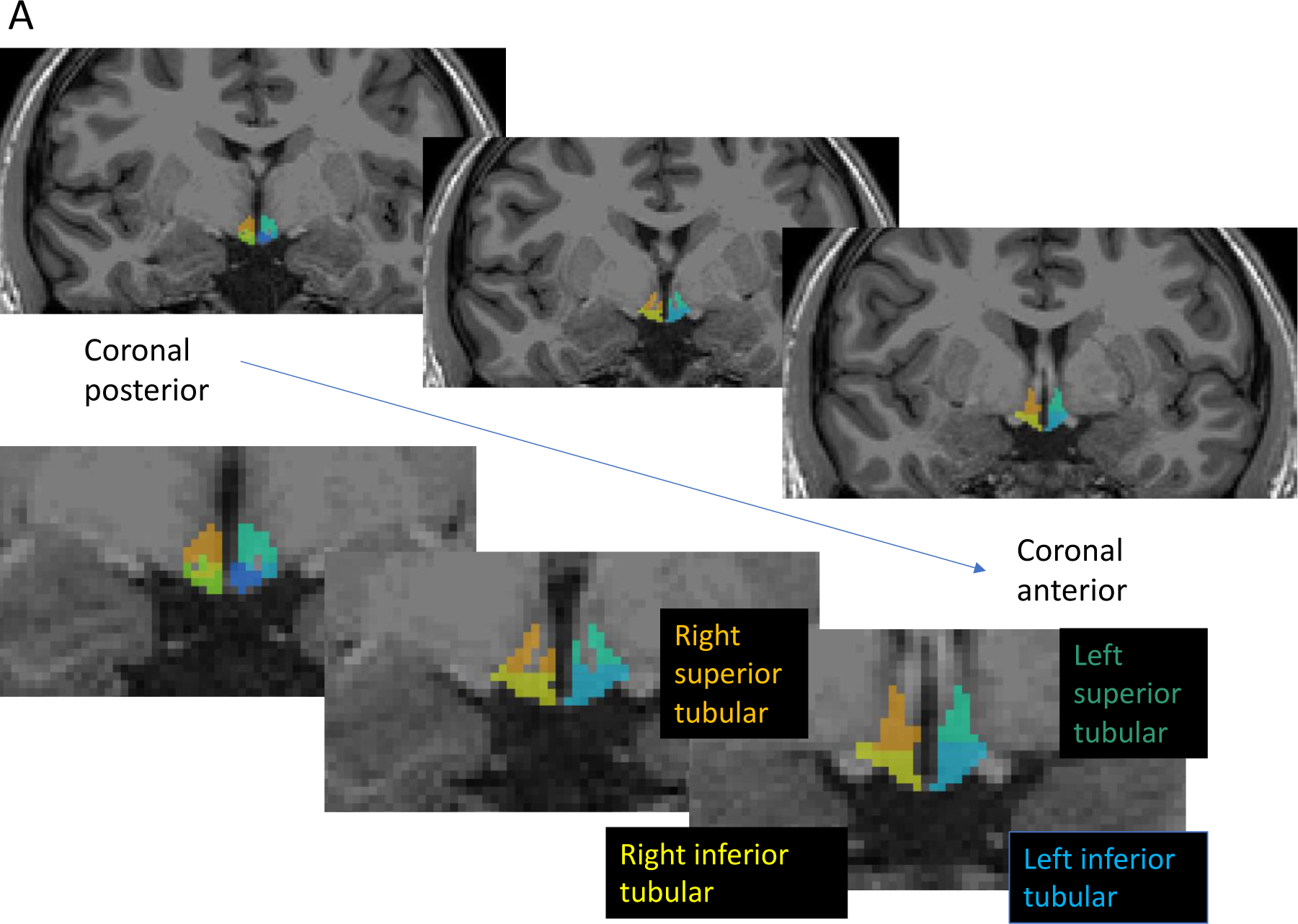
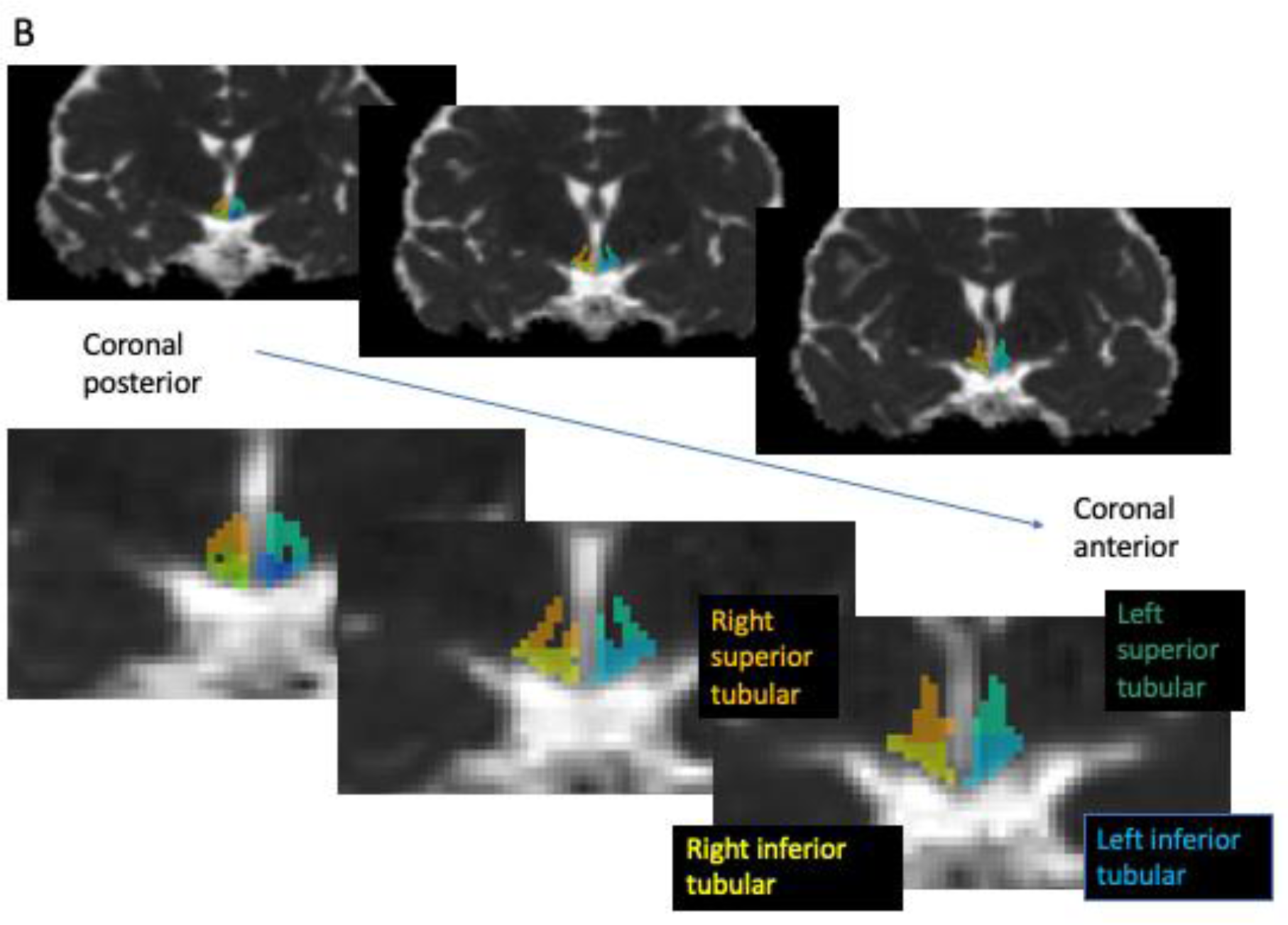
Examples of automatically segmented hypothalamus subnuclei on a participant’s T1-weighted image (A) and respective mean diffusivity (MD) maps (B) in coronal slices. Colors give hypothalamic subnuclei (light blue = left inferior tubular, yellow= right inferior tubular, orange = right superior tubular, turquoise= left superior tubular).

### Hypothalamic MD

To avoid partial volume effects at the border to the adjacent ventricle, we first extracted MD values of the third ventricle based on FS LONG template and the ventricle region according to the Deskian/Killiany atlas (labels 14) (Thomas et al. 2019). Next, we extracted bilateral hippocampus as a control region of non-interest (based on FS LONG automatic segmentation (aseg.mgz) and Deskian/Killiany atlas labels 17 + 53). MD images were coregistered to the respective subject- and time point-specific FS LONG with FSL’s FLIRT using 6 degrees of freedom. Then, the registration matrix was used to coregister the MD images to the anatomical space. Using fslstats (FSL), the mean MD of the third ventricle has been used as an upper threshold for calculating the mean MD of the hypothalamus and the hippocampus, respectively. Analysis code is openly available at https://gitlab.gwdg.de/omega-lab/hypothalamus-segmentation-and-md-extraction.

### Statistical analyses

R version 4.2.2 was used to perform statistical analysis with linear mixed effects models. We controlled for possible confounding effects of body fat mass and BMI since visceral adipose tissue has previously shown to increase proinflammatory cytokines such as TNF-α (Hotamisligil, Shargill, and Spiegelman 1993) and a higher BMI has been linked to increased hypothalamic MD (Thomas et al. 2019). Deviant to the pre-registration, we decided to use lmer function (instead of lm function) in hypotheses 1) and 2) from the R-package lme4 to account for subject as a random factor in order to use all timepoints (up to four per individual) as repeated measures. This required to additionally control for intervention condition and timepoint. Hypotheses 1-4 were tested with the following models:

H1) Null model: lmer(inflammatory_markers∼age+sex+
body_fat_mass+intervention_condition+timepoint+ (1|subj)),
R1: lmer(inflammatory_markers∼fiber_intake+age+sex+ body_fat_mass
+intervention_condition+timepoint+ (1|subj))

H2) Null model: lmer(hypothalamic_MD∼ age+ sex+BMI+intervention_condition+timepoint+
(1|subj))
R1: lmer(hypothalamic_MD∼ fiber_intake+age+ sex+BMI+
intervention_condition+timepoint+ (1|subj))

H3) Null model: lmer(inflammatory_markers∼ intervention_condition+timepoint+ (1|subj))
R1: lmer(inflammatory_ markers∼ timepoint*intervention_condition +
intervention_condition + timepoint + (1|subj))

H4) Null model: lmer(hypothalamic_MD∼ intervention_condition+timepoint+ (1|subj))
R1: lmer(hypothalamic_MD_∼ time point*intervention_condition + intervention_condition +
timepoint + (1|subj)

## Results

### Descriptives

Main analysis included 59 participants with complete baseline assessments (19 women, 40 men) and up to 3 additional assessments adding to maximal 205 observations per outcome (flowchart detailing missing values in **Extended Fig. 1-1**). Participants were 19 to 45 years old (28.3 years ± 6.57 SD), their body fat mass ranged from 7.6% to 39.8% (mean 27.1 % ± 6.6 SD) and self-reported daily habitual fiber intake was diverse and moderate (mean 16.3 g/d ± 6.3 SD, range 1.5 to 30.5) (**Table 2**). Hypothalamic volume and MD values ranged from 707 to 1050 mm^3^ and 0.87*10^−3^ to 1.1*10^−3^ mm²/s, respectively. We observed sex differences in hypothalamic volume size, with higher volumes in males compared to females independent of head size differences. Data across all time points are given in **Extended Fig. 3-1, Extended Table 3-1.**

**Table 2:**
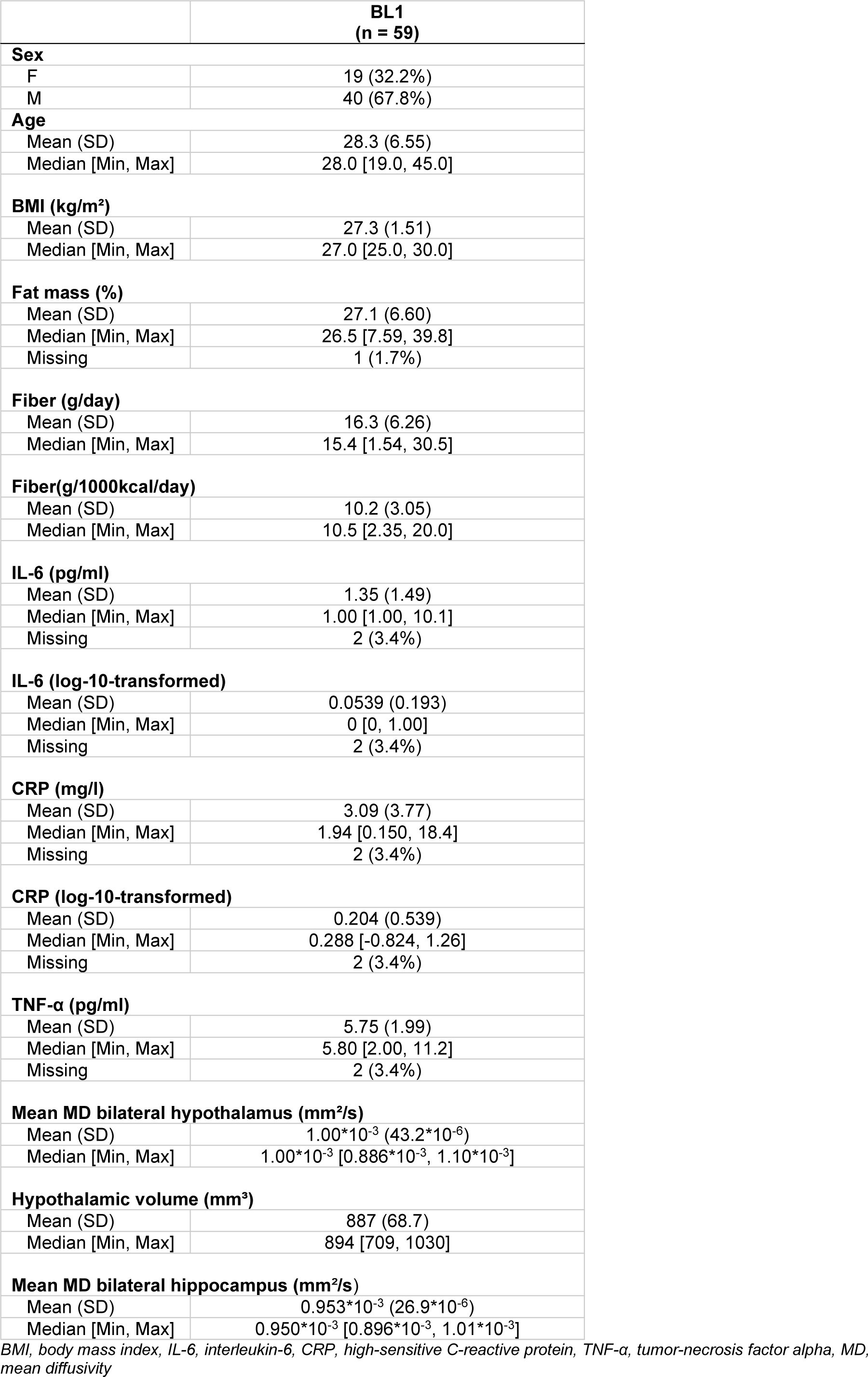
Characteristics of study participants at first assessment (baseline, BL, 1)

As 86% of IL-6 measures laid under the lower limit of quantification we decided to omit IL-6 from statistical analyses (see below for a qualitative evaluation of intervention effects). CRP was used on a log-scale due to skewed distribution (skewness 0.09 for log-transformed data as opposed to 8.18, if not log-transformed).

### Habitual fiber intake, inflammatory markers, and hypothalamic microstructure

As preregistered, we assessed whether habitual dietary fiber intake linked to inflammatory markers and hypothalamic MD. Against our hypotheses, we could not observe significant associations between absolute or relative habitual fiber intake and TNF-α or CRP (model comparisons, all p > 0.37, **Extended Table 3-2 and 3-3**). Neither did we detect significant associations between fiber intake and hypothalamic MD, or hippocampal MD as control region (model comparison, all p > 0.27; **Extended Tables 3-4 and 3-5**).

Notably, male sex predicted lower levels of CRP (ß = −0.6, p < 0.001, Fig. 3A). Additionally, higher body fat (sex-standardized, ß = 0.16, p = 0.002) related to higher CRP levels, independent of the amount of fiber intake (Fig. 3A). This link was not observed for TNF-α (male sex, p = 0.17, body fat, p = 0.22). Moreover, lower hypothalamic MD was borderline associated with higher BMI (Fig. 3B, all p < 0.052**, Extended Table 3-4**).

**Fig. 3:**
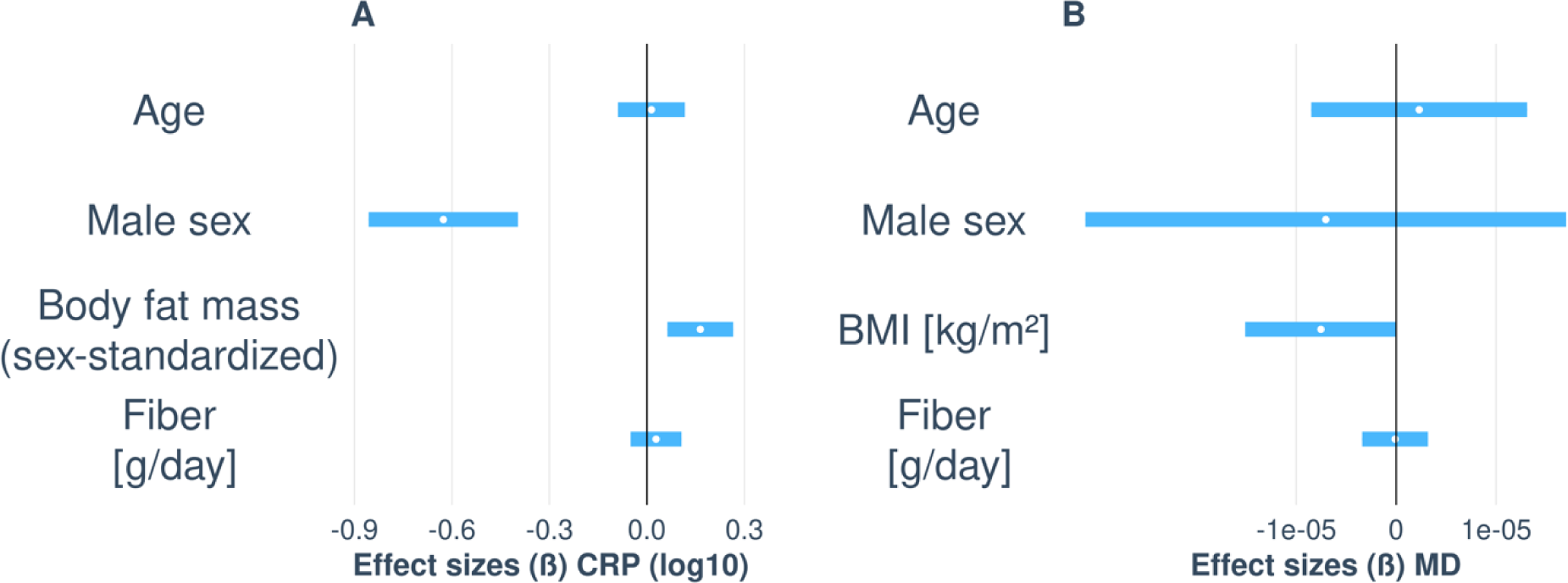
Visualization of unstandardized regression coefficients of baseline models for log-transformed CRP (A) and hypothalamic mean diffusivity (MD) (B) including habitual fiber intake as predictor of interest in comparison to null models (not depicted). Bars depict 95% CI.

Additionally, we explored potential correlations between inflammatory markers and body fat mass in percent stratified by sex due to the observed prediction of lower CRP levels by male sex. Higher body fat somewhat correlated with higher CRP in both females and males, yet the association was not significant in females (Fig. 4; female: r = 0.03, p = 0.74, n = 18; male: ß = 0.18, t = 2.8, p = 0.01, n = 40). We did not observe significant associations with TNF-α and body fat (all p > 0.35).

**Fig. 4:**
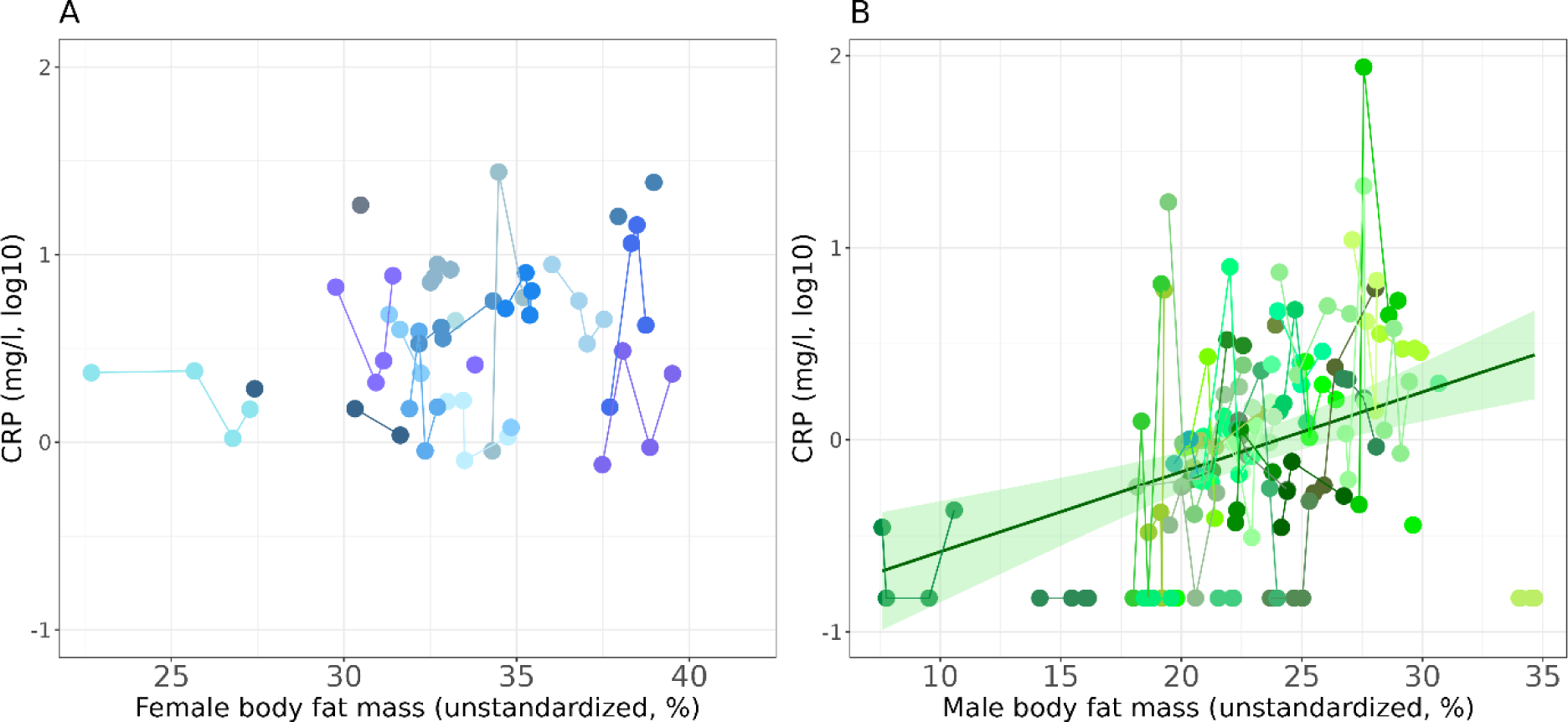
Correlations of body fat mass (FM) in % and log-transformed CRP in females (A) and males (B) Colors code for participant (shading, up to four records) and sex (blue female, green male). Lines indicates regression fit with the lightgreen ribbons represent pointwise 95% CI of the means.

When exploring the borderline association between higher BMI and lower hypothalamic MD stratified for sex, the effect was not evident (Fig. 5; model comparisons, females: p = 0.35, males: p = 0.12).

**Fig.5:**
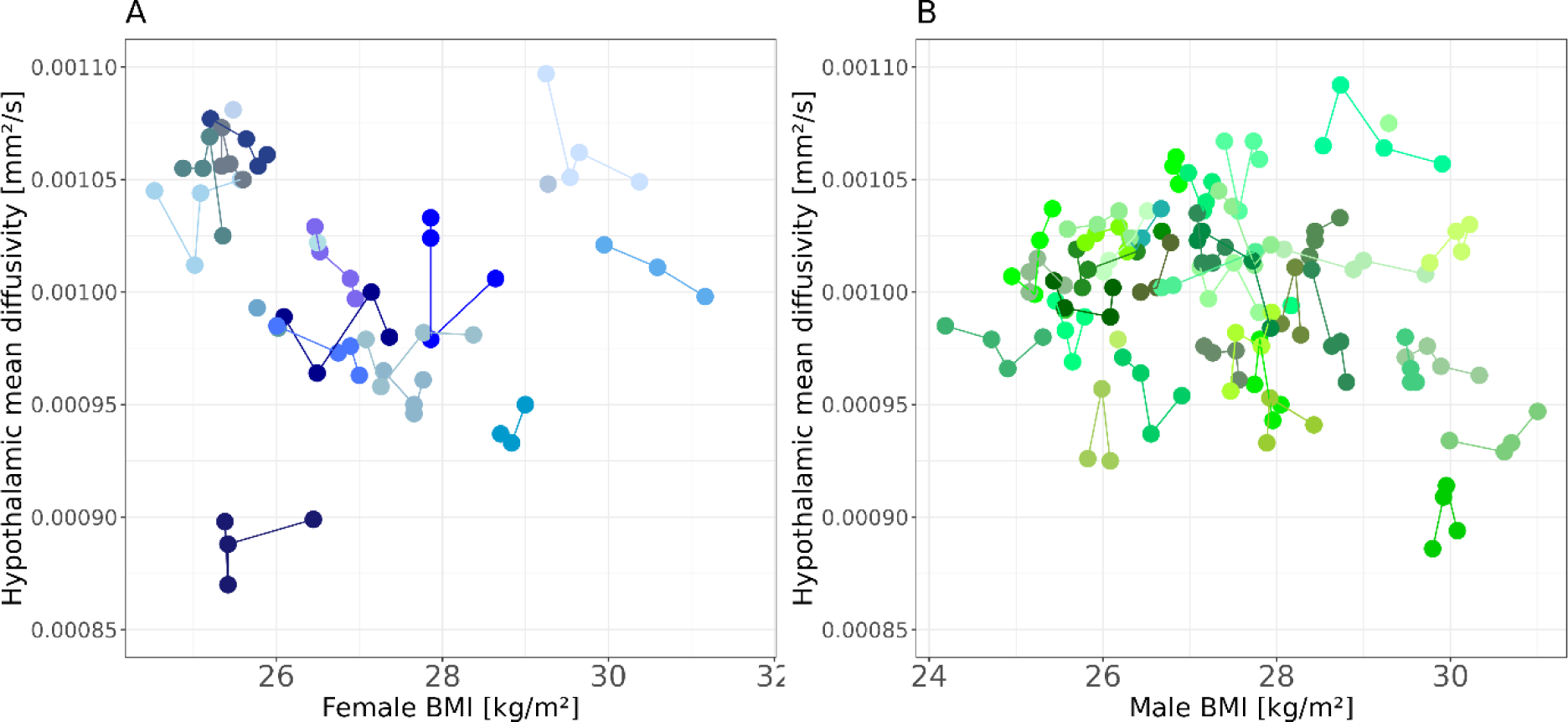
Hypothalamic mean diffusivity (MD) in relation to body mass index (BMI) in females (A) and males (B) Colors code for participant (shading, up to four records) and sex (blue female, green male). Lines connect individuals.

### Effects of 14 days of high-dosed prebiotic fiber intake on inflammatory markers and hypothalamic microstructure

The two-week high-fiber intervention, compared to placebo, did not have significant effects neither on inflammatory markers (Fig. 6A-B, **Extended Table 6-1 and 6-2**, model comparison p = 0.59 for TNF-α, model comparison p = 0.29 for CRP) nor on brain microstructure, i.e., neither hypothalamic MD (Fig. 6C, **Extended Table 6-3,** model comparison p = 0.08) nor the MD of the control region, the hippocampus, were altered by the high-dosed fiber intake (Fig. 6D, **Extended Table 6-4**, model comparison p = 0.87).

**Fig.6:**
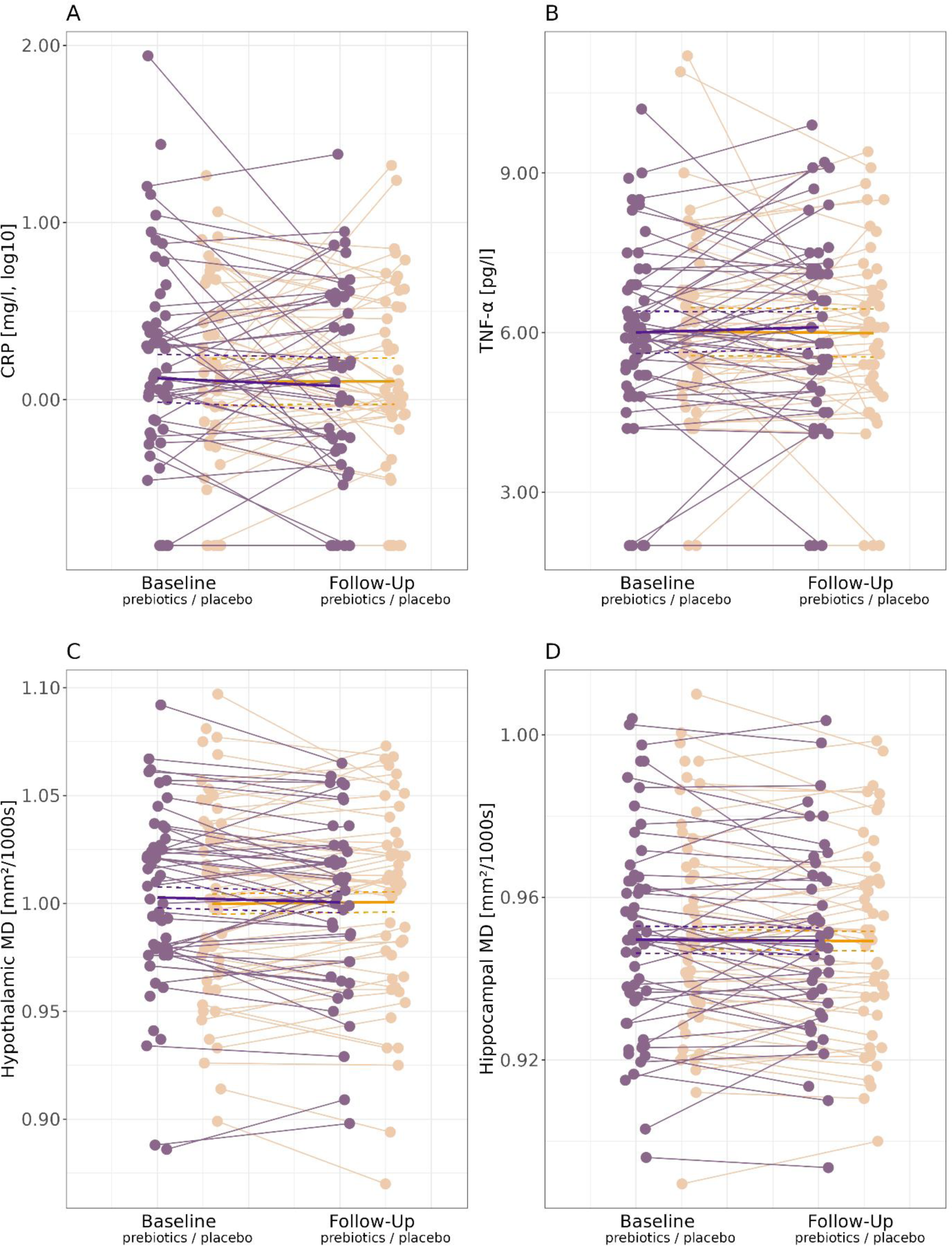
Changes in log-transformed CRP (CRP_log10, A), TNF-a (B), hypothalamic MD (C), and hippocampal MD (D) from baseline (BL) to follow-up (FU) measurements before and after14 days of prebiotic fiber intake (violet) and placebo (light orange), respectively. Lines give individual’s change, bold lines give mean change per condition, dashed lines 95% confidence interval of the mean.

When looking qualitatively at IL-6, when assigning 0 to values with below detection threshold and focusing on participants with two or more measurements, IL-6 measures changed in 8 participants after fiber (5 decreases, 3 increases) and in 11 participants after placebo (7 decreases, 4 increases), while the majority did not change.

In exploratory analysis, we investigated a potential association between peripheral (CRP, TNF-α) and central (hypothalamic MD) inflammatory markers across all four time points. We used linear mixed regression to model the effect of serum inflammatory markers on MD corrected for sex, age, BL/FU visit and intervention as covariates and subject as random factor. Chi-squared tests were used to compare models including the marker against models containing only variables we corrected for. Neither the comparison of log-transformed CRP (p = 0.93) nor TNF-α (p = 0.06) showed a significant effect (Fig. 7, **Extended Tables 7-1 and 7-2**).

**Figure 7:**
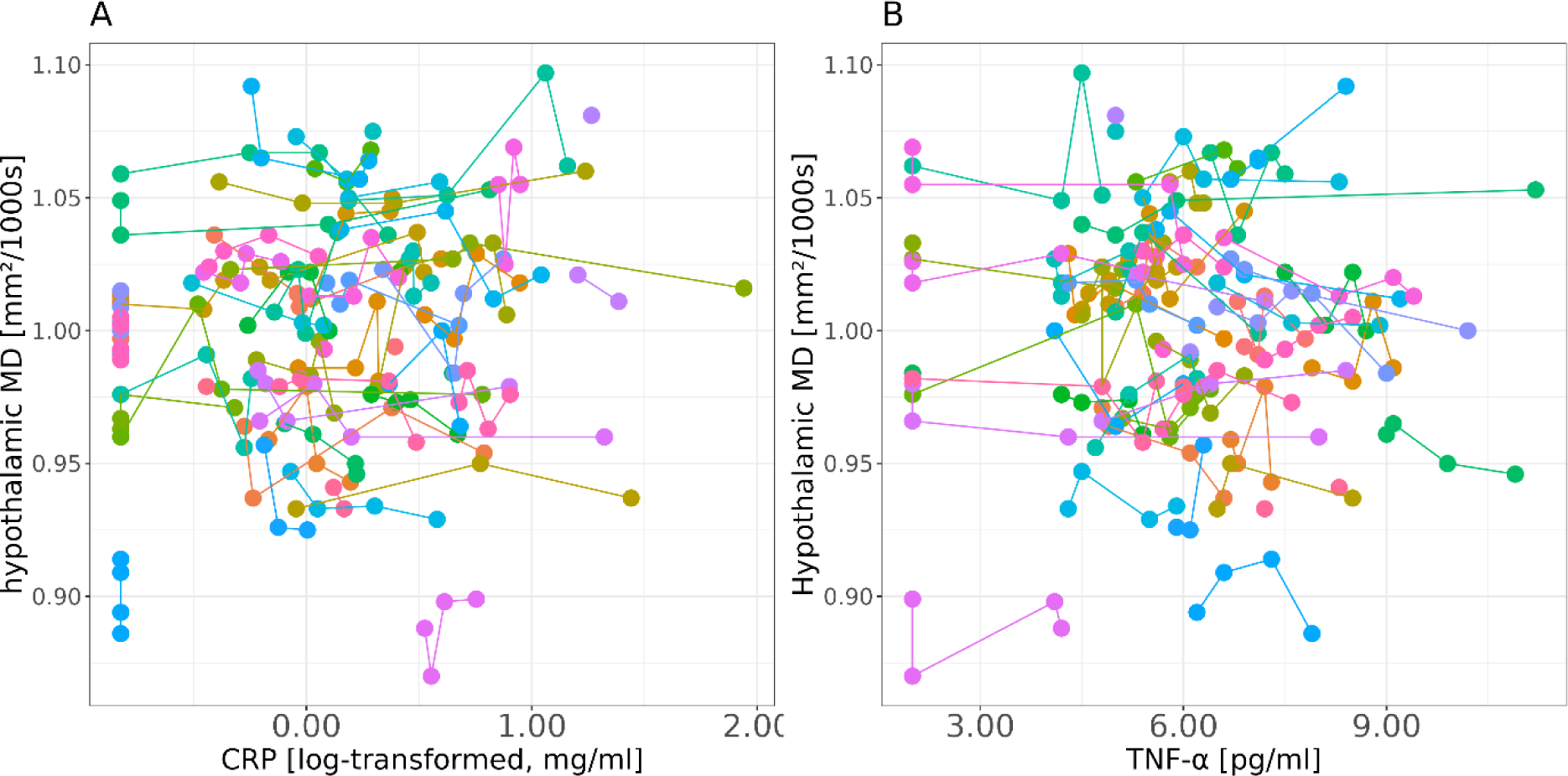
Peripheral compared to central markers of inflammation. (A) log-transformed CRP, (B) TNF-α. Colors code for participants, (up to four records per subject), lines connect participants.

Exploratory analysis of habitual and intervention effects of dietary fiber on hypothalamic volume (corrected for total intracranial volume) did not reveal significant changes (time by group interaction:|ß|= 5.12, p = 0.21, **Extended Tables 7-3 and 7-4**).

## Discussion

In this well-characterized sample of 59 overweight young to middle aged adults, systemic and (neuro-)inflammatory processes measured in blood and hypothalamus were not significantly related to prebiotic fiber intake, i.e. neither to self-reported, habitual intake nor to short-term high-dosed interventional intake for 14 days. Exploratory analyses indicated that sex and body fat related to higher CRP serum levels, while higher BMI was borderline associated with coherence of hypothalamic microstructure, represented in MD.

### Prebiotic fiber intake and peripheral markers of inflammation

Considering blood-based inflammatory markers, we could not confirm that habitual fiber intake related to lower fasting levels of CRP, TNF-α or IL-6, or that a high-fiber intervention decreased these inflammatory markers in our sample. These null results thus question a strong effect of fiber intake on markers of systemic inflammation. Causal evidence for inflammation-lowering effects of high-fiber diets based on interventional studies is indeed scarce: King et al. observed decreases in CRP after three weeks of two different high-fiber diets. However, this observation applied only to a specific subgroup, i.e., the lean group of the study population (17 participants) and not to participants living with obesity (18 participants). Further, there was no placebo control, so that test-retest effects could not be ruled out (King et al. 2007). More recent controlled RCTs observed decreases in TNF-α and IL-6, but not in CRP, in 52 women with type-2 diabetes (mean age 48 years) after 8 weeks of 10 g/d inulin (Dehghan, Pourghassem Gargari, and Asghari Jafar-abadi 2014), and in a group of 42 overweight and obese children after 16 weeks of inulin supplementation (Nicolucci et al. 2017). In general, dietary intervention studies such as ours are often difficult to monitor and can therefore be limited by false reports of habitual dietary intake or insufficient supplement intake. Of note, we did not observe significant effects when controlling for total energy intake to control potential over- or under-reporting, and all participants reported regular supplement intake according to self-report. The lacking effect of fiber intake on systemic inflammation in our study might be also attributed to ceiling effects. For instance, room for inflammatory improvement might have been limited in our study population of overweight young adults as they showed only low-grade inflammation levels (indicated by floor-levels of IL-6) and were metabolically healthy. Participants in our study also already showed an average moderate, habitual dietary fiber intake of 16 g/day, outperforming previous study populations (Dehghan, Pourghassem Gargari, and Asghari Jafar-abadi 2014). In addition, the timeframe of the intervention might have been too short to induce significant effects (two weeks in comparison to 8 (Dehghan, Pourghassem Gargari, and Asghari Jafar-abadi 2014) and 16 weeks (Nicolucci et al. 2017). As CRP-levels were not altered by fiber supplementation in previous studies, it might not be sensitive enough towards fiber diet-induced changes. However, IL-6 levels, which had been lowered in previous studies by fiber supplementation, were in most cases already below detection threshold at baseline in our study population, so already at the healthier end.

On the neural level, hypothalamic microstructure was not related to neither habitual nor supplemental fiber intake. After two weeks of intervention, we observed that mean hypothalamic MD tended to marginally decrease in the fiber intervention condition and marginally increase after placebo intake, however, changes appeared negligible in size and need to be interpreted with caution due to a lack of significance in the time-by-group interaction (p = 0.07). Our results are in line with a previous systematic review reporting rather non-significant effects or very small effect sizes across five RCTs investigating Mediterranean dietary intervention effects on cognition and brain functions (Radd-Vagenas et al. 2018). In a closely controlled environment, however, Song et al. could show that greater adherence to Mediterranean diet, indicative of high-fiber intake, linked to less progression of white matter lesions, probable of vascular and inflammatory pathology, in a prospective study over the course of 5 years (Song et al. 2022). Further, a three-months Mediterranean-DASH intervention led to an increase in surface area of the inferior frontal gyrus suggesting that such a dietary intervention can reverse the potentially adverse effects of previous unhealthy diet on brain structure (Arjmand, Abbas-Zadeh, and Eftekhari 2022). Notably, evidence of diet-body/brain effects might be biased by inaccuracies in study conception and conduction, in particular concerning dietary reporting, underpowered studies and short time-frames (Duplantier and Gardner 2021).

The lack of a significant effect of fiber in our sample might also be explained by the overall healthy condition of participants: hypothalamic MD was lower, indicative of healthier tissue, compared to populations with a three decades-higher mean age (Thomas et al. 2019). Additionally, our study was not powered for the detection of short-term intervention effects on young adults’ brain microstructure. As absence of evidence does not prove evidence of absence, fiber-induced brain changes might be observable in larger or metabolically conspicuous populations. We suggest that future studies should be designed with longer intervention durations or with patient populations (e.g., including aging or metabolic disease). Data pooling can additionally increase sample size to increase the probability of more definite conclusions.

Investigating the effect of confounding variables in exploratory analyses, we observed that higher body fat mass was associated with higher CRP in blood. Detrimental effects of body fat mass on inflammatory markers have been shown previously (Festa et al. 2001). A causal link has been indicated by reversibility experiments in normal-weight, physically active individuals, where weight loss was paralleled by decreased low-grade inflammation (Sarin et al. 2019).

In addition, men had lower levels of CRP similar to pre-midlife populations (Sarin et al. 2019), and higher body fat related to higher CRP in males in our sample. This might have contributed to the observation of lower inflammation values in males. Unfortunately, our sample consisted of more males than females due to more strict exclusion criteria for women. Underlying potential sex differences, the effects of higher BMI on promoting systemic inflammation have previously been shown to be less pronounced in men compared to women (Choi, Joseph, and Pilote 2013). We further found higher BMI to be borderline related to lower hypothalamic MD. Previous results from our and other groups in on average older participants rather indicated that higher BMI, as well as higher age, related to lower MD in the hypothalamus (Birdsill et al. 2017; Dekkers, Jansen, and Lamb 2019; Kullmann et al. 2016; Lampe et al. 2019; Thomas et al. 2019; Sewaybricker et al., 2023), however, one earlier study reported higher values of the apparent diffusion coefficients correlating with higher BMI in the hypothalamus in middle old adults (Alkan et al., 2008). Future studies are needed to further disentangle putative divergent patterns of hypothalamus MD and weight status in different age groups.

Taken together, sex and body composition across the lifespan likely affect peripheral inflammatory markers and brain microstructure more strongly, while dietary habits or short-term dietary interventions exert no strong effects. Indeed, self-reported dietary fiber assessment might have been too coarse, and the two weeks of intervention might not have been long enough to induce changes on markers of systemic inflammation and at the brain microstructural level in this relatively healthy young and healthy population.

### Strengths and Limitations

In sum, limitations of this study include the self-reported dietary measure and short intervention timeframe of two weeks. The validity of self-reported dietary intake and intervention compliance is a central concern of nutrition studies, questioning the overall reliability of (null) findings (Ioannidis 2018). In the current study, we estimated habitual dietary fiber intake from self-report using a validated quantitative food frequency questionnaire (Haftenberger et al. 2010) and developed a detailed diet scoring on macro- and micronutrient level (Thieleking et al. 2023). Our double-blinded interventional, cross-over within-subject design providing >200 repeated measures can be rated as gold standard. Supplementary fiber intake compliance during intervention was high according to daily diaries (average missed intakes 1.25 ± 1.8, min = 0, max = 9). Within our study, we also collected stool samples which showed significant shifts in the gut microbiome after fiber intervention (Medawar 2021, Medawar, in press) indicating high compliance with the supplementation. In sum, quality of the dietary data and intervention compliance in the present analysis can be considered reliable.

Regarding population characteristics, we selected intuitive, omnivorous dieters in an overweight range. Therefore, the BMI range covered only a part of different body constitutions, so that our results cannot be transferred to other weight categories. Nonetheless, we deeply phenotyped all participants at each study time point collecting various serum markers, questionnaire data and high-resolution brain microstructural and structural data. Thereby, we reduced biases possibly interfering with effects of interest, namely by adding confounding factors and random effects to our statistical models. For the difficult extraction of hypothalamic volume and MD values due to proximity to the third ventricle, we followed a state-of-art processing pipeline resulting in reliable measures of the hypothalamus. We also controlled for known influences on brain microstructure. We additionally want to emphasize that prior to completion of data acquisition and data analysis, we preregistered a detailed analysis plan (https://osf.io/uzbav) and provided open code for re-use.

## Conclusion

In this study, we investigated whether a habitual high-fiber diet and a two-week prebiotic high-fiber intervention would reduce inflammatory processes. More precisely, we hypothesized a decrease in blood levels of CRP and TNF-α and in hypothalamic MD. Our findings do not support evidence for habitual or supplemented dietary fiber acting in an anti-inflammatory manner in this young, overweight population. Rather, sex and body composition were of higher importance for prediction of peripheral inflammation. Future trials are needed that implement more diverse age and weight status groups and a longer timeframe for prebiotic fiber supplementation, to advance our understanding of diet-brain modifications.

## Data availability

Scripts for hypothalamus segmentation and extraction of MD are made available here: https://gitlab.gwdg.de/omega-lab/hypothalamus-MD.

## Author Contributions

Study conception: EM, RT, AV, AVW, MS; data collection: EM, ET, RT; data curation: EM, ET; brain imaging data processing: EM, FB; first manuscript draft: EM, ET; all authors contributed to and accepted the final draft.

## Data Availability

Data availability
Scripts for hypothalamus segmentation and extraction of MD are made available here:
https://gitlab.gwdg.de/omega-lab/hypothalamus-MD.

## Acknowledgements

We thank all participants of the GUT-BRAIN study. This work was funded by grants of the German Research Foundation (DFG), contract grant number 209933838 CRC1052-03 A1 to A.V.W and M.S., and by the Berlin School of Mind and Brain (stipend for E.M.) and the German Foundation for Environment (stipend for E.M.). The inulin supplement was sponsored by the manufacturer BENEO GmbH, Mannheim, Germany. We would like to thank all those who contributed to the conduction of the GUT-BRAIN study, whether it was recruitment of participants, data acquisition, data organization, data storage or data curation and analysis: Larissa de Biasi, Anne-Katrin Brecht, Anna Bujanow, Leonie Disch, Lina Eisenberg, Silke Friedrich, Laura Hesse, Niklas Hlubek, Bettina Johst, Mandy Jochemko, Anke Kummer, Domenica Klank, Lorenz Lemcke, Ramona Menger, Lynn Mosesku, Maria Pärisch, Susan Prejawa, Lukas Recker, Lennard Schneidewind, Emira Shehabi, Hannah Stock, Torsten Schlumm, Christian Schneider, Anna-Luisa Wehle, Charlotte Wiegank, and Marie Zedler.

