## Supplementary Information for "Habitual and supplemented prebiotic diets and their links to inflammatory serum markers and hypothalamic microstructure in young, overweight adults: a pre-registered study"

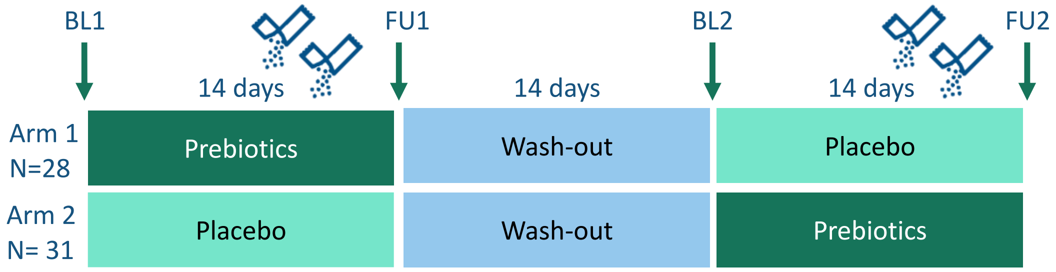


**Figure 1: Study design.** Each participant underwent up to four assessments: Baseline 1 (BL1, before first intervention), Follow-up 1 (FU1, after first intervention), Baseline 2 (before second intervention), Follow-up 2 (after second intervention). In-between two assessments participants supplemented their diet either with "prebiotics" (30g of inulin) or "placebo" (isocaloric maltodextrin). Participants were randomly allocated to study arm 1 or 2 which determined the order of supplement intake. A washout period of at least 14 days was set between interventions.

**Table 1: Diffusion-weighted imaging (DWI) Quality Control.** Group-wise quality metrics provided by eddy squad for DWI data.

| | Signal-to-noise ratio (SNR) | Contrast-to-noise ratio (CNR) | avg. absolute motion [mm] | avg. relative motion [mm] |
|---|---|---|---|---|
| Mean | 40.93 | 4.13 | 0.27 | 0.12 |
| SD | 7.15 | 0.73 | 0.19 | 0.06 |
| Mean +/- 1 SD | 33.78 | 3.40 | 0.46 | 0.17 |
| Mean +/- 2 SD | 26.64 | 2.67 | 0.65 | 0.23 |

Overall, data quality of DWI data was judged as very good without extreme values according to quality metrics provided by eddy squad.

Hypothalamus segmentation.

Deviant to the pre-registration, we used automated segmentation of the bilateral hypothalamus using a deep learning algorithm (Billot et al. 2020) implemented in python scripts (v3.6) due to faster and more reliable results. Briefly, four hypothalamic regions were segmented at each hemisphere (**Figure 2**). Visual checks for correct segmentation on the structural image were done for all subjects. Volumes for total bilateral hypothalamus were extracted for each individual and for each of the up to four datapoints. Subnuclei were disregarded for statistical inference analysis to reduce Type II errors.

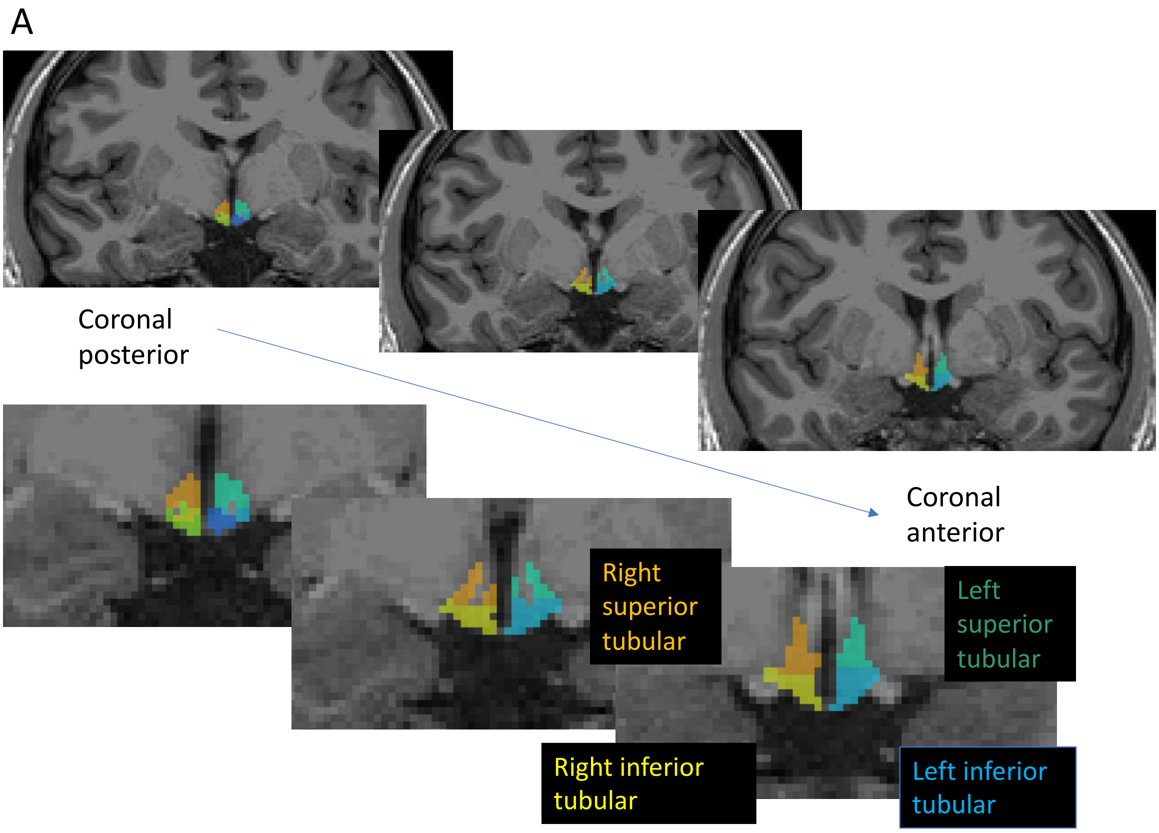

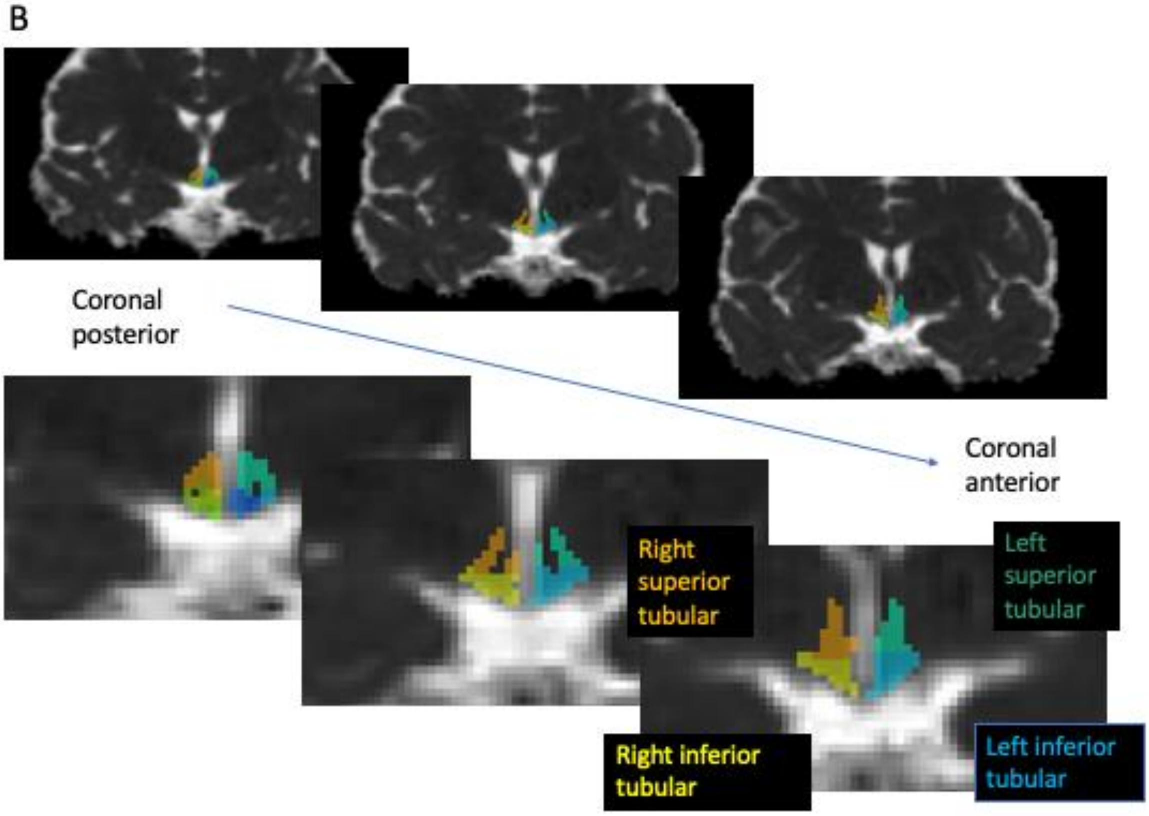


**Fig. 2: Examples of automatically segmented hypothalamus subnuclei on a participant's T1-weighted image (A) and respective mean diffusivity (MD) maps (B) in coronal slices**. Colors give hypothalamic subnuclei (light blue = left inferior tubular, yellow= right inferior tubular, orange = right superior tubular, turquoise= left superior tubular).

**Table 2: Characteristics of study participants at first assessment (baseline, BL, 1).**

| | BL1 (n = 59) |
|---|---|
| **Sex** | |
| F | 19 (32.2%) |
| M | 40 (67.8%) |
| **Age** | |
| Mean (SD) | 28.3 (6.55) |
| Median [Min, Max] | 28.0 [19.0, 45.0] |
| **BMI (kg/m²)** | |
| Mean (SD) | 27.3 (1.51) |
| Median [Min, Max] | 27.0 [25.0, 30.0] |
| **Fat mass (%)** | |
| Mean (SD) | 27.1 (6.60) |
| Median [Min, Max] | 26.5 [7.59, 39.8] |
| Missing | 1 (1.7%) |
| **Fiber (g/day)** | |
| Mean (SD) | 16.3 (6.26) |
| Median [Min, Max] | 15.4 [1.54, 30.5] |
| **Fiber(g/1000kcal/day)** | |
| Mean (SD) | 10.2 (3.05) |
| Median [Min, Max] | 10.5 [2.35, 20.0] |
| **IL-6 (pg/ml)** | |
| Mean (SD) | 1.35 (1.49) |
| Median [Min, Max] | 1.00 [1.00, 10.1] |
| Missing | 2 (3.4%) |
| **IL-6 (log-10-transformed)** | |
| Mean (SD) | 0.0539 (0.193) |
| Median [Min, Max] | 0 [0, 1.00] |
| Missing | 2 (3.4%) |
| **CRP (mg/l)** | |
| Mean (SD) | 3.09 (3.77) |
| Median [Min, Max] | 1.94 [0.150, 18.4] |
| Missing | 2 (3.4%) |
| **CRP (log-10-transformed)** | |
| Mean (SD) | 0.204 (0.539) |
| Median [Min, Max] | 0.288 [-0.824, 1.26] |
| Missing | 2 (3.4%) |
| **TNF-α (pg/ml)** | |
| Mean (SD) | 5.75 (1.99) |
| Median [Min, Max] | 5.80 [2.00, 11.2] |
| Missing | 2 (3.4%) |
| **Mean MD bilateral hypothalamus (mm²/s)** | |
| Mean (SD) | $1.00*10^{-3}$ ($43.2*10^{-6}$) |
| Median [Min, Max] | $1.00*10^{-3}$ [$0.886*10^{-3}$, $1.10*10^{-3}$] |
| **Hypothalamic volume (mm³)** | |
| Mean (SD) | 887 (68.7) |
| Median [Min, Max] | 894 [709, 1030] |
| **Mean MD bilateral hippocampus (mm²/s)** | |
| Mean (SD) | $0.953*10^{-3}$ ($26.9*10^{-6}$) |
| Median [Min, Max] | $0.950*10^{-3}$ [$0.896*10^{-3}$, $1.01*10^{-3}$] |

*BMI, body mass index, IL-6, interleukin-6, CRP, high-sensitive C-reactive protein, TNF-α, tumor-necrosis factor alpha, MD, mean diffusivity*

**Habitual fiber intake, inflammatory markers, and hypothalamic microstructure**

As preregistered, we assessed whether habitual dietary fiber intake linked to inflammatory markers and hypothalamic MD. Against our hypotheses, we could not observe significant associations between absolute or relative habitual fiber intake and TNF-α or CRP (model comparisons, all $p > 0.37$, **Extended Table 3-2 and 3-3**). Neither did we detect significant associations between fiber intake and hypothalamic MD, or hippocampal MD as control region (model comparison, all $p > 0.27$; **Extended Tables 3-4 and 3-5**).

Notably, male sex predicted lower levels of CRP (ß = -0.6, $p < 0.001$, **Fig. 3A**). Additionally, higher body fat (sex-standardized, ß = 0.16, $p = 0.002$) related to higher CRP levels, independent of the amount of fiber intake (**Fig. 3A**). This link was not observed for TNF-α (male sex, $p = 0.17$, body fat, $p = 0.22$). Moreover, lower hypothalamic MD was borderline associated with higher BMI (**Fig. 3B**, all $p < 0.052$, **Extended Table 3-4**).

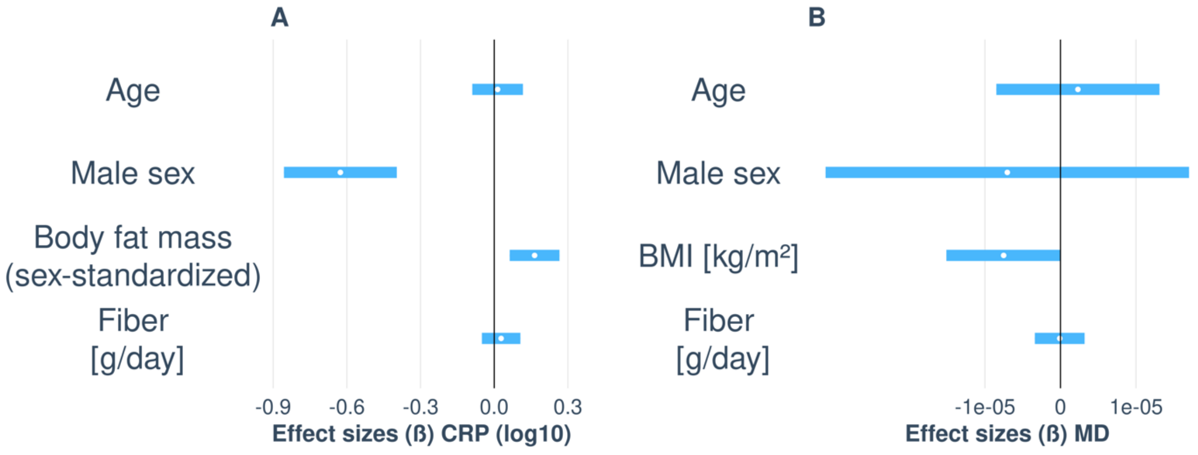


**Fig. 3: Visualization of unstandardized regression coefficients of baseline models for log-transformed CRP (A) and hypothalamic mean diffusivity (MD) (B),** including habitual fiber intake as predictor of interest in comparison to null models (not depicted). Bars depict 95% CI.

Additionally, we explored potential correlations between inflammatory markers and body fat mass in percent stratified by sex due to the observed prediction of lower CRP levels by male sex. Higher body fat somewhat correlated with higher CRP in both females and males, yet the association was not significant in females (**Fig. 4;** female: r = 0.03, p = 0.74, n = 18; male: ß = 0.18, t = 2.8, p = 0.01, n = 40). We did not observe significant associations with TNF-α and body fat (all p > 0.35).

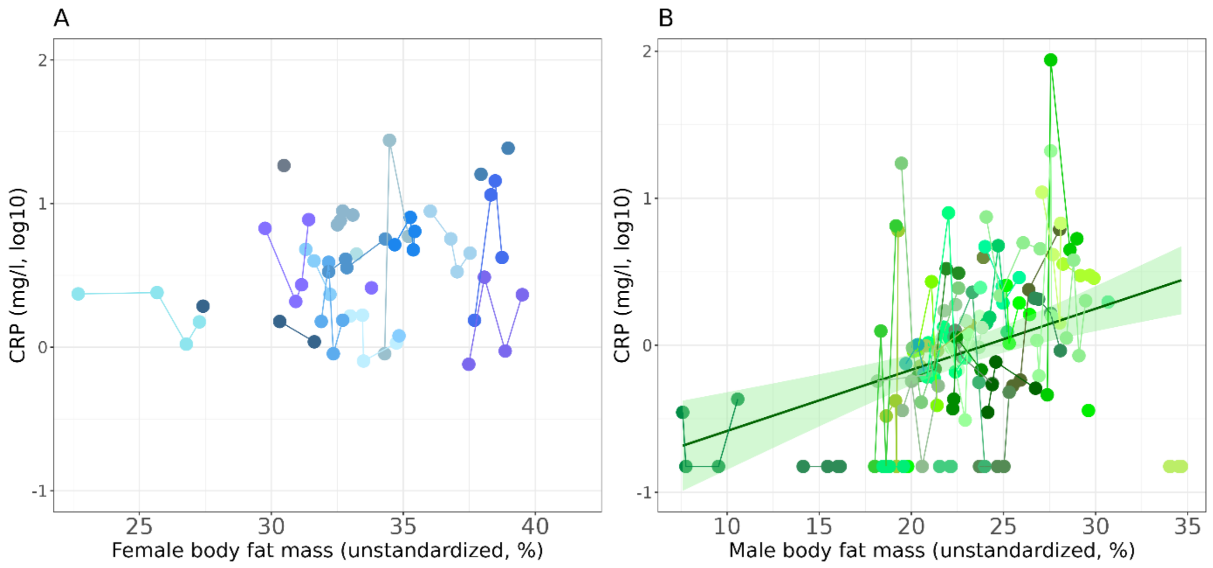


**Fig. 4**: Correlations of body fat mass (FM) in % and log-transformed CRP in females (A) and males (B). Colors code for participant (shading, up to four records) and sex (blue female, green male). Lines indicates regression fit with the lightgreen ribbons represent pointwise 95% CI of the means.

When exploring the borderline association between higher BMI and lower hypothalamic MD stratified for sex, the effect was not evident (**Fig. 5**; model comparisons, females: p = 0.35, males: p = 0.12).

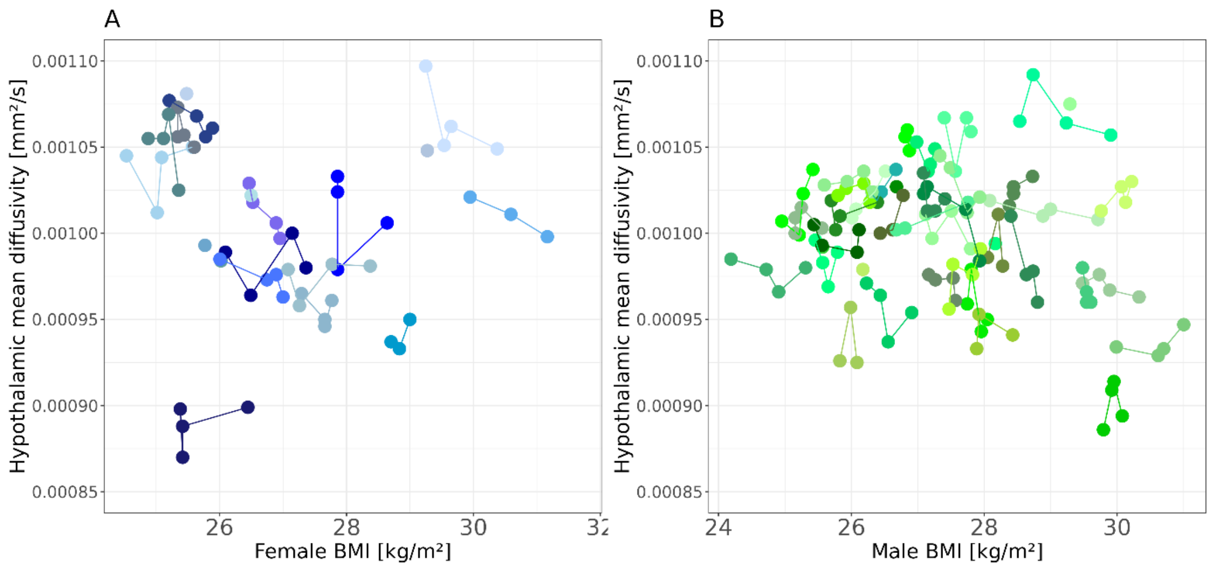


**Fig.5: Hypothalamic mean diffusivity (MD) in relation to body mass index (BMI) in females (A) and males (B).** Colors code for participant (shading, up to four records) and sex (blue female, green male). Lines connect individuals.

#### Effects of 14 days of high-dosed prebiotic fiber intake on inflammatory markers and hypothalamic microstructure

The two-week high-fiber intervention, compared to placebo, did not have significant effects neither on inflammatory markers (**Fig. 6A-B**, **Extended Table 6-1 and 6-2**, model comparison $p = 0.59$ for TNF-α, model comparison $p = 0.29$ for CRP) nor on brain microstructure, i.e., neither hypothalamic MD (**Fig. 6C**, **Extended Table 6-3,** model comparison $p = 0.08$) nor the MD of the control region, the hippocampus, were altered by the high-dosed fiber intake (**Fig. 6D**, **Extended Table 6-4**, model comparison $p = 0.87$).

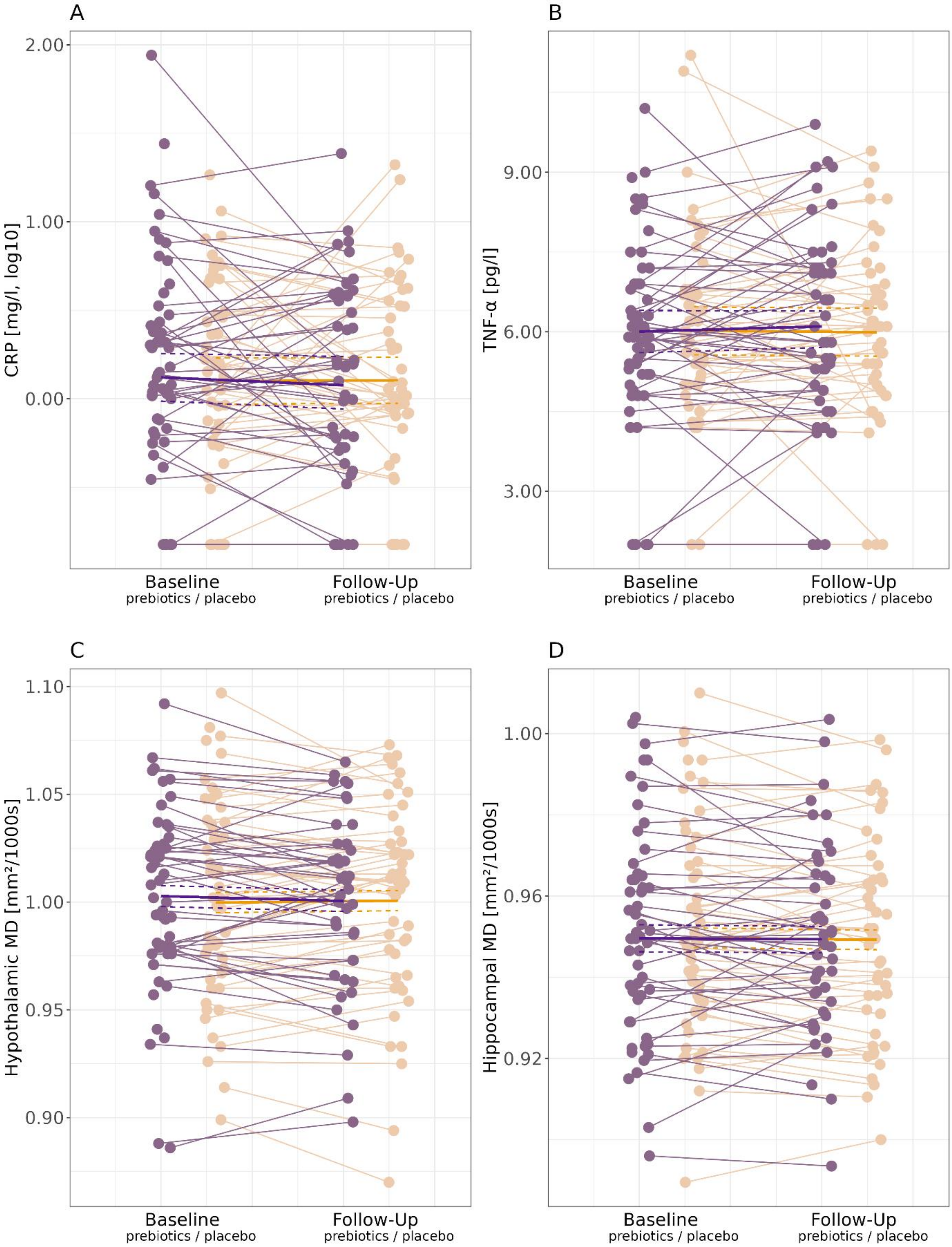


**Fig.6: Changes in log-transformed CRP (CRP_log10, A), TNF-a (B), hypothalamic MD (C), and hippocampal MD (D) from baseline (BL) to follow-up (FU) measurements** before and after14 days of prebiotic fiber intake (violet) and placebo (light orange), respectively. Lines give individual's change, bold lines give mean change per condition, dashed lines 95% confidence interval of the mean.

When looking qualitatively at IL-6, when assigning 0 to values with below detection threshold and focusing on participants with two or more measurements, IL-6 measures changed in 8 participants after fiber (5 decreases, 3 increases) and in 11 participants after placebo (7 decreases, 4 increases), while the majority did not change.

In exploratory analysis, we investigated a potential association between peripheral (CRP, TNF-α) and central (hypothalamic MD) inflammatory markers across all four time points. We used linear mixed regression to model the effect of serum inflammatory markers on MD corrected for sex, age, BL/FU visit and intervention as covariates and subject as random factor. Chi-squared tests were used to compare models including the marker against models containing only variables we corrected for. Neither the comparison of log-transformed CRP ($p = 0.93$) nor TNF- α ($p = 0.06$) showed a significant effect (**Fig. 7, Extended Tables 7-1 and 7-2**).

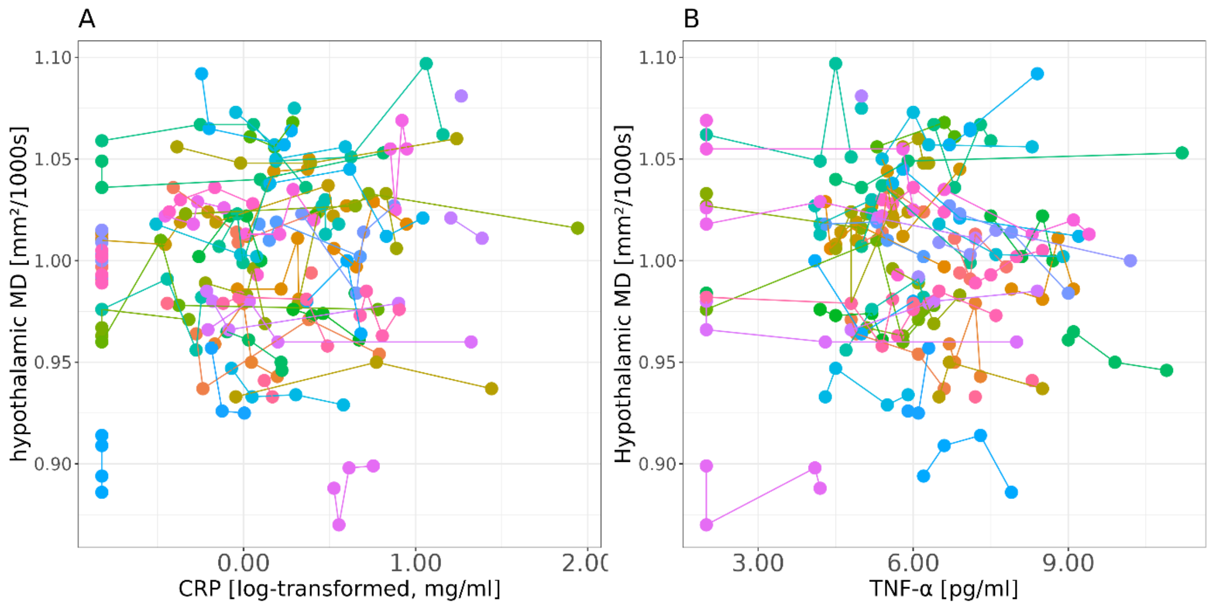


**Figure 7: Peripheral compared to central markers of inflammation.** (A) log-transformed CRP, (B) TNF-α. Colors code for participants, (up to four records per subject), lines connect participants.

Exploratory analysis of habitual and intervention effects of dietary fiber on hypothalamic volume (corrected for total intracranial volume) did not reveal significant changes (time by group interaction: $|\beta| = 5.12$, $p = 0.21$, **Extended Tables 7-3 and 7-4**).

Medawar E, Beyer F, Thieleking R, Haange SB, Rolle-Kampczyk U, Reinicke M, Chakaroun R, von Bergen M, Stumvoll M, Villringer A, Witte AV (in press) A prebiotic diet changes neural correlates of food decision-making in overweight adults: a randomized controlled within-subject cross-over trial. Gut.

Morrison, Douglas J., and Tom Preston. 2016. “Formation of Short Chain Fatty Acids by the Gut Microbiota and Their Impact on Human Metabolism.” *Gut Microbes* 7(3): 189–200. http://dx.doi.org/10.1080/19490976.2015.1134082.

Nicolucci, Alissa C. et al. 2017. “Prebiotics Reduce Body Fat and Alter Intestinal Microbiota in Children Who Are Overweight or With Obesity.” *Gastroenterology* 153(3): 711–22. http://dx.doi.org/10.1053/j.gastro.2017.05.055.

Radd-Vagenas, Sue et al. 2018. “Effect of the Mediterranean Diet on Cognition and Brain Morphology and Function: A Systematic Review of Randomized Controlled Trials.” *Am J Clin Nutr* 107: 389–404. http://www.crd.york.ac.uk/PROSPERO/display_record.asp?ID=.

Rocha, D. M. et al. 2016. “Saturated Fatty Acids Trigger TLR4-Mediated Inflammatory Response.” *Atherosclerosis* 244: 211–15. http://dx.doi.org/10.1016/j.atherosclerosis.2015.11.015.

Sarin, H. V. et al. 2019. “Substantial Fat Mass Loss Reduces Low-Grade Inflammation and Induces Positive Alteration in Cardiometabolic Factors in Normal-Weight Individuals.” *Scientific Reports* 9(1): 1–14.

De Silva, Akila, and Stephen R. Bloom. 2012. “Gut Hormones and Appetite Control: A Focus on PYY and GLP-1 as Therapeutic Targets in Obesity.” *Gut and Liver* 6(1): 10–20.

Sewaybricker LE, Huang A, Chandrasekaran S, Melhorn SJ, Schur EA. The Significance of Hypothalamic Inflammation and Gliosis for the Pathogenesis of Obesity in Humans. Endocr Rev. 2023 Mar 4;44(2):281-296. doi: 10.1210/endrev/bnac023.

Song, Suhang et al. 2022. “Mediterranean Diet and White Matter Hyperintensity Change over Time in Cognitively Intact Adults.” https://doi.org/10.3390/nu14173664.

Thaler, Joshua P, and Michael W Schwartz. 2010. “Minireview: Inflammation and Obesity Pathogenesis: The Hypothalamus Heats Up.” *Endocrinology* 151(9): 4109–15. https://pubmed.ncbi.nlm.nih.gov/20573720.

Thieleking, Ronja, Lennard Schneidewind, Arsene Kanyamibwa, and Hendrik Hartmann. 2023. “Nutrient Scoring for the DEGS1 - FFQ – from Food Intake to Nutrient Intake.” *BMC Nutrition*: 1–16. https://doi.org/10.1186/s40795-022-00636-2.

Thomas, K. et al. 2019. “Higher Body Mass Index Is Linked to Altered Hypothalamic Microstructure.” *Scientific Reports* 9(1): 1–11.

Wannamethee, S. Goya, Peter H. Whincup, Mary C. Thomas, and Naveed Sattar. 2009. “Associations between Dietary Fiber and Inflammation, Hepatic Function, and Risk of Type 2 Diabetes in Older Men: Potential Mechanisms for the Benefits of Fiber on Diabetes Risk.” *Diabetes Care* 32(10): 1823–25.

### Supplementary Material

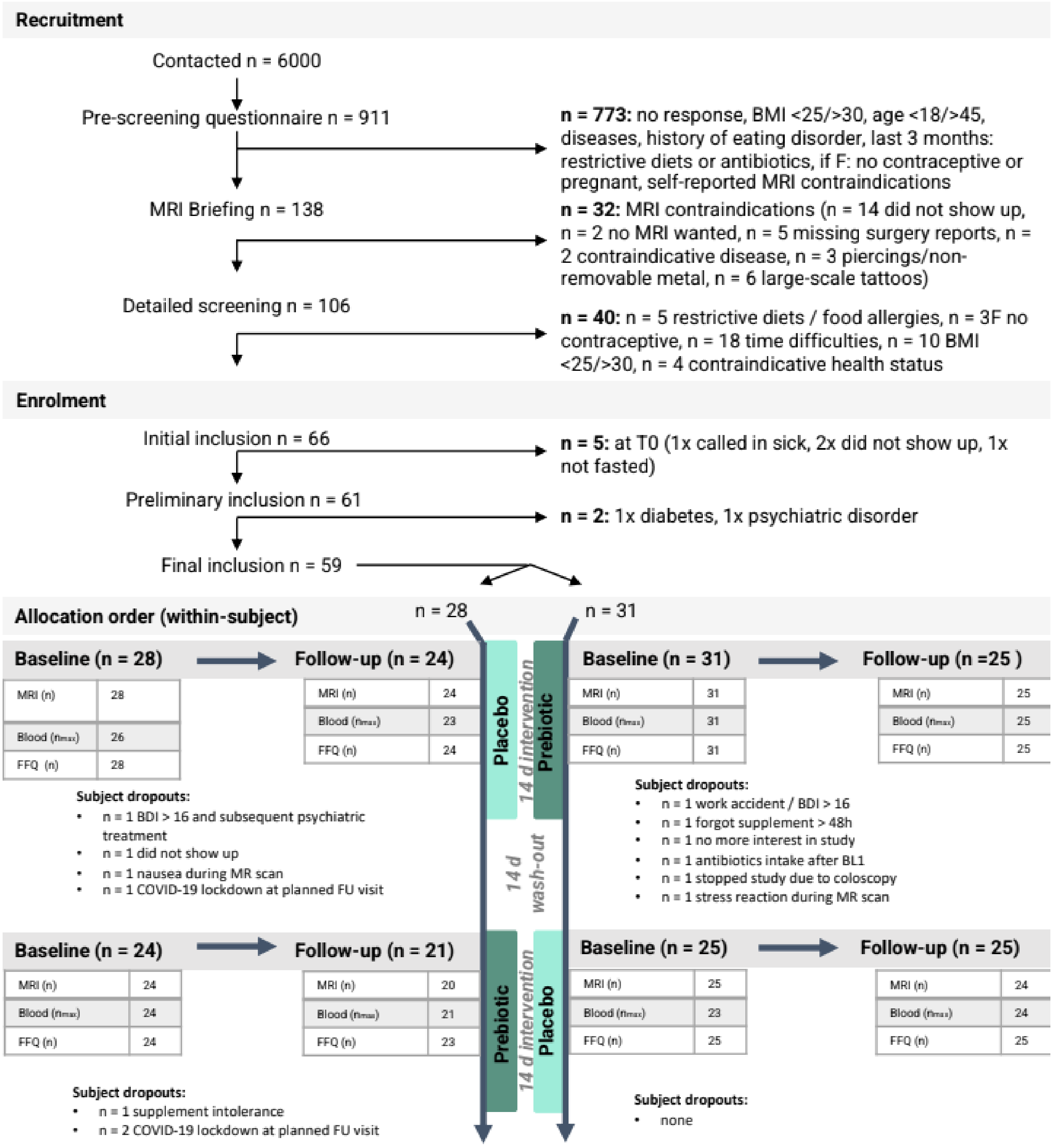


**Extended Fig. 1-1:** Flowchart showing numbers and reasons of participants´ dropouts

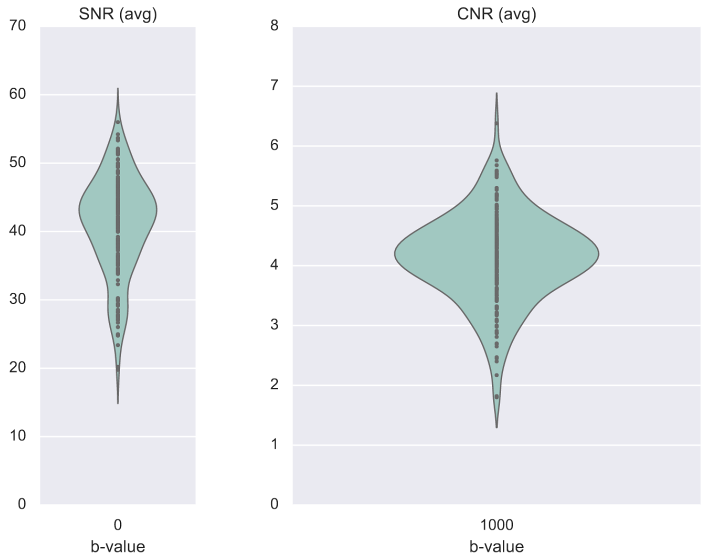


**Extended Fig. 2-1: Violin plot for quality assessment of diffusion-weighted images.** Depicted are the distributions of signal-to-noise ratios (SNR) for b=0 images (without diffusion gradient) and contrast-to-noise ratios (CNR) for images with b=1000.

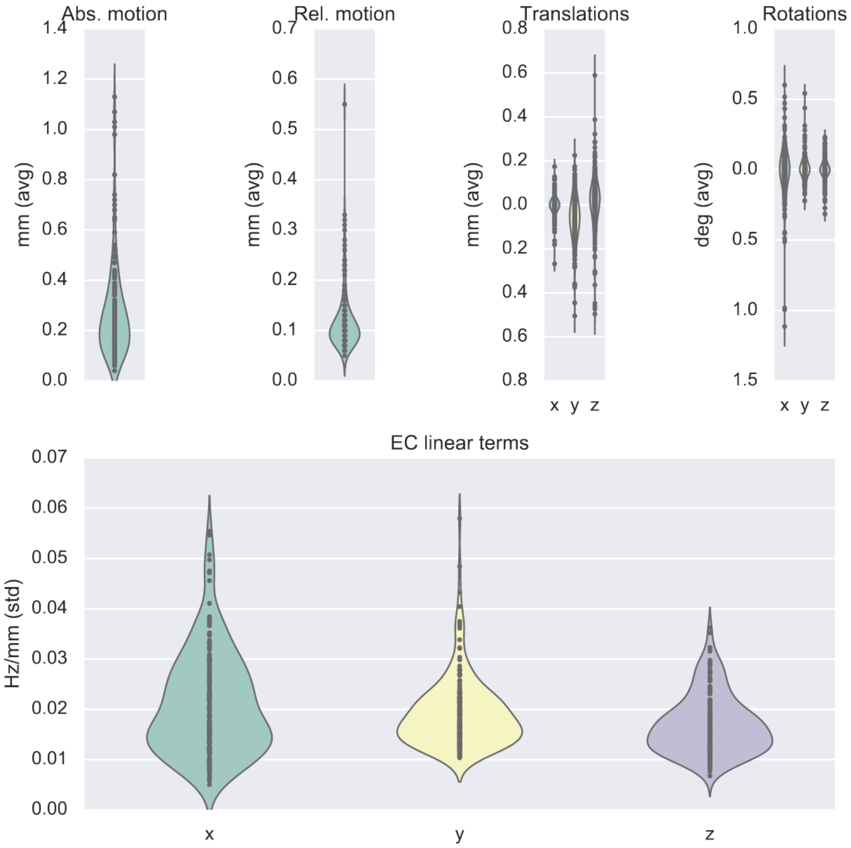


**Extended Fig. 2-2: Group-wise quality metrics provided by eddy squad for diffusion-weighted images.**

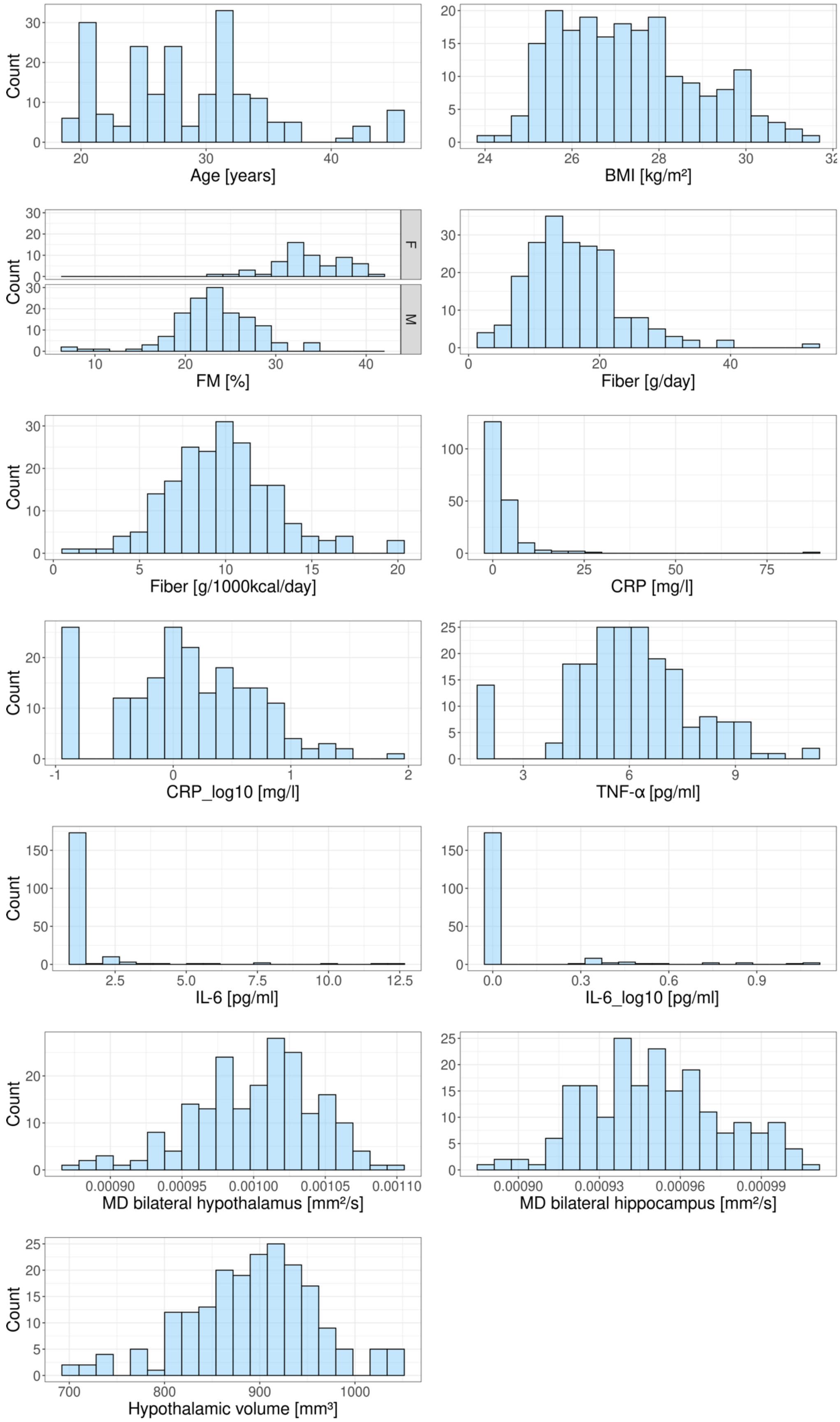
Count
Age [years]
BMI [kg/m²]
F
M
FM [%]
Fiber [g/day]
Fiber [g/1000kcal/day]
CRP [mg/l]
CRP_log10 [mg/l]
TNF-α [pg/ml]
IL-6 [pg/ml]
IL-6_log10 [pg/ml]
MD bilateral hypothalamus [mm²/s]
MD bilateral hippocampus [mm²/s]
Hypothalamic volume [mm³]

**Extended Fig. 3-1:** Histograms of all Variables of Interest and Covariates.

**Extended Table 3-1**: Descriptive table of variables of interest and covariates across all time points. Note that Fiber and Placebo intervention followed randomized order.

| | **Inulin (Fiber)** | | **Maltodextrin (Placebo)** | |
|---|---|---|---|---|
| | **BL (N=55)** | **FU (N=48)** | **BL (N=53)** | **FU (N=49)** |
| **Sex** | | | | |
| F | 17 (30.9%) | 14 (29.2%) | 16 (30.2%) | 14 (28.6%) |
| M | 38 (69.1%) | 34 (70.8%) | 37 (69.8%) | 35 (71.4%) |
| **Age** | | | | |
| Mean (SD) | 28.7 (6.37) | 28.5 (6.24) | 28.1 (6.38) | 28.5 (6.17) |
| Median [Min, Max] | 28.0 [19.0, 45.0] | 27.5 [19.0, 45.0] | 27.0 [19.0, 45.0] | 28.0 [19.0, 45.0] |
| **BMI (kg/m²)** | | | | |
| Mean (SD) | 27.2 (1.50) | 27.4 (1.64) | 27.4 (1.61) | 27.3 (1.67) |
| Median [Min, Max] | 27.1 [24.5, 30.2] | 27.2 [24.2, 30.6] | 27.3 [25.0, 31.2] | 27.1 [24.9, 31.7] |
| Missing | 0 (0%) | 2 (4.2%) | 0 (0%) | 0 (0%) |
| **Fat mass (%)** | | | | |
| Mean (SD) | 26.2 (6.49) | 26.2 (6.30) | 27.0 (6.66) | 26.0 (6.46) |
| Median [Min, Max] | 24.8 [7.59, 38.5] | 25.0 [10.6, 39.0] | 26.7 [9.53, 41.6] | 25.2 [7.76, 38.9] |
| Missing | 0 (0%) | 2 (4.2%) | 1 (1.9%) | 0 (0%) |
| **Fiber (g/day)** | | | | |
| Mean (SD) | 16.1 (6.52) | 16.4 (7.64) | 16.2 (7.11) | 15.8 (7.54) |
| Median [Min, Max] | 14.8 [1.54, 31.6] | 15.7 [2.12, 51.2] | 15.4 [6.48, 40.0] | 14.1 [2.18, 39.3] |
| **Fiber(g/1000kcal/day)** | | | | |
| Mean (SD) | 9.88 (2.68) | 9.60 (3.05) | 9.88 (3.37) | 9.83 (3.62) |
| Median [Min,Max] | 10.2 [2.35, 16.1] | 9.28 [4.29, 20.3] | 9.87 [3.15, 20.0] | 9.48 [1.41, 19.9] |
| **IL-6 (pg/ml)** | | | | |
| Mean (SD) | 1.56 (2.08) | 1.11 (0.363) | 1.35 (1.10) | 1.52 (1.86) |
| Median [Min, Max] | 1.00 [1.00, 12.0] | 1.00 [1.00, 2.30] | 1.00 [1.00, 7.50] | 1.00 [1.00, 12.2] |
| Missing | 0 (0%) | 2 (4.2%) | 4 (7.5%) | 2 (4.1%) |
| **IL-6 (log-10-transformed)** | | | | |
| Mean | 0.0752 (0.239) | 0.0310 (0.102) | 0.0707 (0.188) | 0.0788 (0.229) |

| | | | | |
|---|---|---|---|---|
| (SD) | | | | |
| Median [Min,Max] | 0 [0, 1.08] | 0 [0, 0.362] | 0 [0, 0.875] | 0 [0, 1.09] |
| Missing | 0 (0%) | 2 (4.2%) | 4 (7.5%) | 2 (4.1%) |
| **CRP (mg/l)** | | | | |
| Mean (SD) | 4.66 (12.3) | 2.61 (3.97) | 2.69 (3.43) | 2.64 (4.00) |
| Median [Min, Max] | 1.41 [0.150, 87.3] | 1.39 [0.150, 24.3] | 1.50 [0.150, 18.4] | 1.04 [0.150, 21.0] |
| Missing | 0 (0%) | 2 (4.2%) | 4 (7.5%) | 2 (4.1%) |
| **CRP (log-10-transformed)** | | | | |
| Mean (SD) | 0.186 (0.607) | 0.0764 (0.572) | 0.119 (0.562) | 0.104 (0.538) |
| Median [Min, Max] | 0.149 [-0.824, 1.94] | 0.140 [-0.824, 1.39] | 0.176 [-0.824, 1.26] | 0.0170 [-0.824, 1.32] |
| Missing | 0 (0%) | 2 (4.2%) | 4 (7.5%) | 2 (4.1%) |
| **TNF-α (pg/ml)** | | | | |
| Mean (SD) | 5.94 (1.79) | 6.10 (1.85) | 5.94 (1.85) | 5.99 (1.67) |
| Median [Min, Max] | 6.00 [2.00, 10.2] | 5.95 [2.00, 9.90] | 5.80 [2.00, 11.2] | 6.10 [2.00, 9.40] |
| Missing | 0 (0%) | 2 (4.2%) | 4 (7.5%) | 2 (4.1%) |
| **Mean MD bilateral hypothalamus (mm²/s)** | | | | |
| Mean (SD) | $1.00*10^{-3}$ ($41.1*10^{-6}$) | $1.00*10^{-3}$ ($39.4*10^{-6}$) | $1.00*10^{-3}$ ($45.6*10^{-6}$) | $1.00e*10^{-3}$ ($45.5*10^{-6}$) |
| Median [Min, Max] | $1.00*10^{-3}$ [$0.89*10^{-3}$, $1.09*10^{-3}$] | $1.01*10^{-3}$ [$0.90*10^{-3}$, $1.07*10^{-3}$] | $1.00*10^{-3}$ [$0.90*10^{-3}$, $1.10*10^{-3}$] | $1.01*10^{-3}$ [$0.87*10^{-3}$, $1.07*10^{-3}$] |
| Missing | 0 (0%) | 3 (6.3%) | 0 (0%) | 1 (2.0%) |
| **Hypothalamic volume (mm³)** | | | | |
| Mean (SD) | 893 (63.8) | 899 (62.6) | 887 (73.5) | 893 (69.6) |
| Median [Min, Max] | 900 [743, 1030] | 908 [707, 1040] | 894 [709, 1050] | 893 [720, 1040] |
| Missing | 0 (0%) | 3 (6.3%) | 0 (0%) | 1 (2.0%) |
| **Mean MD bilateral hippocampus (mm²/s)** | | | | |
| Mean (SD) | $0.951*10^{-3}$ ($25.5*10^{-6}$) | $0.949*10^{-3}$ ($23.3*10^{-6}$) | $0.952*10^{-3}$ ($25.9*10^{-6}$) | $0.949*10^{-3}$ ($24.2*10^{-6}$) |
| Median [Min, Max] | $0.950*10^{-3}$ [$0.896*10^{-3}$, $1.00*10^{-3}$] | $0.948*10^{-3}$ [$0.894*10^{-3}$, $1.00*10^{-3}$] | $0.949*10^{-3}$ [$0.890*10^{-3}$, $1.01*10^{-3}$] | $0.950*10^{-3}$ [$0.900*10^{-3}$, $0.999*10^{-3}$] |
| Missing | 0 (0%) | 3 (6.3%) | 0 (0%) | 1 (2.0%) |

*BMI, body mass index, IL-6, interleukin-6, CRP, high-sensitive C-reactive protein, TNF-α, tumor-necrosis factor alpha, MD, mean diffusivity*

#### **Extended Table 3-2a:** Models for TNF-α, habitual fiber intake effect

| Model | Response | Fixed effects | Random effects |
|---|---|---|---|
| 1 | TNF-α | age+sex+body fat mass, sex-standardized+time+intervention | 1 \| subject |
| 1.1 | TNF-α | fiber per gramm+age+sex+body fat mass, sex-standardized+ time+ intervention | 1 \| subject |
| 1.2 | TNF-α | fiber per 1000kcal+age+sex+body fat mass, sex-standardized+ time+intervention | 1 \| subject |

#### **Extended Table 3-2b:** Coefficients of Model 1

| | Estimate | Std. Error | df | t value | Pr(>\|t\|) |
|---|---|---|---|---|---|
| intercept | 3.948 | 0.890 | 60.5 | 4.435 | 0.000 |
| age | 0.053 | 0.031 | 57.6 | 1.709 | 0.093 |
| sex (male) | 0.591 | 0.427 | 56.6 | 1.383 | 0.172 |
| body fat mass, sex-standardized | -0.230 | 0.185 | 72.5 | -1.246 | 0.217 |
| time (follow-up) | 0.072 | 0.173 | 143.4 | 0.416 | 0.678 |
| intervention (fiber) | 0.092 | 0.173 | 144.4 | 0.529 | 0.598 |

#### **Extended Table 3-2c:** Coefficients of Model 1.1

| | Estimate | Std. Error | df | t value | Pr(>\|t\|) |
|---|---|---|---|---|---|
| intercept | 3.962 | 0.910 | 64.7 | 4.355 | 0.000 |
| fiber per gramm | -0.002 | 0.020 | 188.3 | -0.080 | 0.936 |
| age | 0.053 | 0.031 | 57.7 | 1.702 | 0.094 |
| sex (male) | 0.595 | 0.431 | 57.4 | 1.380 | 0.173 |
| body fat mass, sex-standardized | -0.233 | 0.190 | 80.6 | -1.231 | 0.222 |
| time (follow-up) | 0.072 | 0.173 | 142.9 | 0.416 | 0.678 |
| intervention (fiber) | 0.092 | 0.174 | 143.9 | 0.530 | 0.597 |

#### **Extended Table 3-2d:** Coefficients of Model 1.2

| | Estimate | Std. Error | df | t value | Pr(>\|t\|) |
|---|---|---|---|---|---|
| intercept | 3.678 | 0.943 | 71.3 | 3.901 | 0.000 |
| fiber per 1000kcal | 0.036 | 0.041 | 187.2 | 0.878 | 0.381 |
| age | 0.049 | 0.031 | 59.2 | 1.548 | 0.127 |
| sex (male) | 0.639 | 0.431 | 57.7 | 1.482 | 0.144 |
| body fat mass, sex-standardized | -0.212 | 0.186 | 75.3 | -1.138 | 0.259 |

| | Estimate | Std. Error | df | t value | Pr(>\|t\|) |
|---|---|---|---|---|---|
| time (follow-up) | 0.075 | 0.173 | 142.7 | 0.434 | 0.665 |
| intervention (fiber) | 0.104 | 0.174 | 144.1 | 0.597 | 0.552 |

**Extended Table 3-2e:** Model comparison (ANOVA) of Model 1 and Model 1.1

| | npar | AIC | BIC | logLik | deviance | Chisq | Df | Pr(>Chisq) |
|---|---|---|---|---|---|---|---|---|
| fit1 | 8 | 721 | 747 | -353 | 705 | NA | NA | NA |
| fit1.1 | 9 | 723 | 753 | -353 | 705 | 0.008 | 1 | 0.927 |

**Extended Table 3-2f:** Model comparison (ANOVA) of Model 1 and Model 1.2

| | npar | AIC | BIC | logLik | deviance | Chisq | Df | Pr(>Chisq) |
|---|---|---|---|---|---|---|---|---|
| fit1 | 8 | 721 | 747 | -353 | 705 | NA | NA | NA |
| fit1.2 | 9 | 722 | 752 | -352 | 704 | 0.793 | 1 | 0.373 |

**Extended Table 3-3a:** Models for CRP, habitual fiber intake effect

| Model | Response | Fixed effects | Random effects |
|---|---|---|---|
| 3 | CRP | age+sex+body fat mass, sex-standardized+time+ intervention | 1 \| subject |
| 3.1 | CRP | fiber per gramm+age+sex+body fat mass, sex-standardized+time+intervention | 1 \| subject |
| 3.2 | CRP | fiber per 1000kcal+age+sex+body fat mass, sex-standardized+time+ intervention | 1 \| subject |

**Extended Table 3-3b:** Coefficients of Model 3

| | Estimate | Std. Error | df | t value | Pr(>\|t\|) |
|---|---|---|---|---|---|
| intercept | 0.512 | 0.241 | 57.8 | 2.123 | 0.038 |
| age | 0.003 | 0.008 | 54.2 | 0.359 | 0.721 |
| sex (male) | -0.616 | 0.115 | 53.1 | -5.344 | 0.000 |
| body fat mass, sex-standardized | 0.158 | 0.050 | 64.1 | 3.129 | 0.003 |
| time (follow-up) | -0.047 | 0.052 | 141.5 | -0.890 | 0.375 |
| intervention (fiber) | 0.000 | 0.052 | 142.6 | -0.004 | 0.997 |

**Extended Table 3-3c:** Coefficients of Model 3.1

| | Estimate | Std. Error | df | t value | Pr(>\|t\|) |
|---|---|---|---|---|---|
| intercept | 0.476 | 0.249 | 61.9 | 1.912 | 0.061 |
| fiber per gramm | 0.004 | 0.006 | 183.9 | 0.706 | 0.481 |
| age | 0.002 | 0.008 | 53.3 | 0.264 | 0.793 |

| | Estimate | Std. Error | df | t value | Pr(>\|t\|) |
|---|---|---|---|---|---|
| sex (male) | -0.626 | 0.117 | 53.2 | -5.343 | 0.000 |
| body fat mass, sex-standardized | 0.167 | 0.052 | 71.9 | 3.179 | 0.002 |
| time (follow-up) | -0.047 | 0.052 | 140.2 | -0.903 | 0.368 |
| intervention (fiber) | -0.001 | 0.052 | 141.4 | -0.021 | 0.983 |

**Extended Table 3-3d:** Coefficients of Model 3.2

| | Estimate | Std. Error | df | t value | Pr(>\|t\|) |
|---|---|---|---|---|---|
| intercept | 0.450 | 0.259 | 69.4 | 1.735 | 0.087 |
| fiber per 1000kcal | 0.008 | 0.012 | 189.0 | 0.695 | 0.488 |
| age | 0.002 | 0.009 | 55.4 | 0.232 | 0.817 |
| sex (male) | -0.605 | 0.117 | 54.2 | -5.167 | 0.000 |
| body fat mass, sex-standardized | 0.162 | 0.051 | 66.8 | 3.176 | 0.002 |
| time (follow-up) | -0.046 | 0.052 | 140.5 | -0.876 | 0.382 |
| intervention (fiber) | 0.003 | 0.053 | 142.1 | 0.049 | 0.961 |

**Extended Table 3-3e:** Model comparison (ANOVA) of Model 3 and Model 3.1

| | npar | AIC | BIC | logLik | deviance | Chisq | Df | Pr(>Chisq) |
|---|---|---|---|---|---|---|---|---|
| fit3 | 8 | 241 | 267 | -112 | 225 | NA | NA | NA |
| fit3.1 | 9 | 242 | 272 | -112 | 224 | 0.463 | 1 | 0.496 |

**Extended Table 3-3f:** Model comparison (ANOVA) of Model 3 and Model 3.2

| | npar | AIC | BIC | logLik | deviance | Chisq | Df | Pr(>Chisq) |
|---|---|---|---|---|---|---|---|---|
| fit3 | 8 | 241 | 267 | -112 | 225 | NA | NA | NA |
| fit3.2 | 9 | 242 | 272 | -112 | 224 | 0.47 | 1 | 0.493 |

**Extended Table 3-4a:** Hypothalamic MD, habitual fiber intake effect

| Model | Response | Fixed effects | Random effects |
|---|---|---|---|
| 5 | MD_hyp | age+sex+BMI+time+ intervention | 1 \| subject |
| 5.1 | MD_hyp | fiber per gramm+age+sex+BMI+time+ intervention | 1 \| subject |
| 5.2 | MD_hyp | fiber per 1000kcal+age+sex+BMI+ time+intervention | 1 \| subject |

**Extended Table 3-4b:** Coefficients of Model 5

| | Estimate | Std. Error | df | t value | Pr(>\|t\|) |
|---|---|---|---|---|---|
| intercept | 0.0011274 | $72.8*10^{-6}$ | 4885373 | 15.475 | 0.000 |

| | Estimate | Std. Error | df | t value | Pr(>\|t\|) |
|---|---|---|---|---|---|
| age | 0.0000004 | $0.9*10^{-7}$ | 4885373 | 0.417 | 0.677 |
| sex (male) | -0.0000071 | $12.3*10^{-6}$ | 4885373 | -0.576 | 0.564 |
| BMI | -0.0000048 | $2.4*10^{-6}$ | 4885373 | -1.953 | 0.051 |
| time (follow-up) | -0.0000015 | $1.8*10^{-6}$ | 4885373 | -0.833 | 0.405 |
| interventionplacebo | 0.0000012 | $1.8*10^{-6}$ | 4885373 | 0.640 | 0.522 |

#### **Extended Table 3-4c:** Coefficients of Model 5.1

| | Estimate | Std. Error | df | t value | Pr(>\|t\|) |
|---|---|---|---|---|---|
| intercept | 0.0011277 | $73.4*10^{-6}$ | $3.72*10^{8}$ | 15.368 | 0.000 |
| fiber per gramm | 0.0000000 | $0.3*10^{-6}$ | $3.72*10^{8}$ | -0.058 | 0.954 |
| age | 0.0000004 | $0.9*10^{-6}$ | $3.72*10^{8}$ | 0.419 | 0.675 |
| sex (male) | -0.0000070 | $12.3*10^{-6}$ | $3.72*10^{8}$ | -0.573 | 0.566 |
| BMI | -0.0000048 | $2.5*10^{-6}$ | $3.72*10^{8}$ | -1.946 | 0.052 |
| time (follow-up) | -0.0000015 | $1.9*10^{-6}$ | $3.72*10^{8}$ | -0.832 | 0.405 |
| interventionplacebo | 0.0000012 | $1.8*10^{-6}$ | $3.72*10^{8}$ | 0.639 | 0.523 |

#### **Extended Table 3-4d:** Coefficients of Model 5.2

| | Estimate | Std. Error | df | t value | Pr(>\|t\|) |
|---|---|---|---|---|---|
| intercept | 0.0011412 | $73.9*10^{-6}$ | 2494473 | 15.433 | 0.000 |
| fiber per 1000kcal | -0.0000005 | $0.5*10^{-6}$ | 2494473 | -1.089 | 0.276 |
| age | 0.0000004 | $0.9*10^{-6}$ | 2494473 | 0.470 | 0.638 |
| sex (male) | -0.0000076 | $12.3*10^{-6}$ | 2494473 | -0.620 | 0.536 |
| BMI | -0.0000051 | $2.5*10^{-6}$ | 2494473 | -2.081 | 0.037 |
| time (follow-up) | -0.0000016 | $1.8*10^{-6}$ | 2494473 | -0.874 | 0.382 |
| interventionplacebo | 0.0000013 | $1.8*10^{-6}$ | 2494473 | 0.728 | 0.467 |

#### **Extended Table 3-4e:** Model comparison (ANOVA) of Model 5 and Model 5.1

| | npar | AIC | BIC | logLik | deviance | Chisq | Df | Pr(>Chisq) |
|---|---|---|---|---|---|---|---|---|
| fit5 | 8 | -3707 | -3681 | 1862 | -3723 | NA | NA | NA |
| fit5.1 | 9 | -3705 | -3676 | 1862 | -3723 | 0.003 | 1 | 0.955 |

#### **Extended Table 3-4f:** Model comparison (ANOVA) of Model 5 and Model 5.2

| | npar | AIC | BIC | logLik | deviance | Chisq | Df | Pr(>Chisq) |
|---|---|---|---|---|---|---|---|---|
| fit5 | 8 | -3707 | -3681 | 1862 | -3723 | NA | NA | NA |
| fit5.2 | 9 | -3707 | -3677 | 1862 | -3725 | 1.2 | 1 | 0.273 |

#### **Extended Table 3-5a:** Hippocampal MD, habitual fiber intake effect

| Model | Response | Fixed effects | Random effects |
|---|---|---|---|
| 7 | MD_hippo | age+sex+BMI+time+ intervention | 1 \| subject |
| 7.1 | MD_hippo | fiber per gramm+age+sex+body fat mass, sex-standardized+ time+intervention | 1 \| subject |
| 7.2 | MD_hippo | fiber per 1000kcal+age+sex+body fat mass, sex-standardized+ time+ intervention | 1 \| subject |

#### **Extended Table 3-5b:** Coefficients of Model 7

| | Estimate | Std. Error | df | t value | Pr(>\|t\|) |
|---|---|---|---|---|---|
| intercept | 0.0010224 | $41.2*10^{-6}$ | 20.8 | 24.835 | 0.000 |
| age | -0.0000003 | $0.5*10^{-6}$ | 20.1 | -0.501 | 0.622 |
| sex (male) | 0.0000038 | $7.5*10^{-6}$ | 20.1 | 0.504 | 0.620 |
| BMI | -0.0000024 | $1.4*10^{-6}$ | 20.4 | -1.745 | 0.096 |
| time (follow-up) | -0.0000010 | $1.0*10^{-6}$ | 17.7 | -1.046 | 0.310 |
| intervention (fiber) | -0.0000001 | $1.0*10^{-6}$ | 17.7 | -0.104 | 0.918 |

#### **Extended Table 3-5c:** Coefficients of Model 7.1

| | Estimate | Std. Error | df | t value | Pr(>\|t\|) |
|---|---|---|---|---|---|
| intercept | 0.0010166 | $41.7*10^{-6}$ | 20.6 | 24.378 | 0.000 |
| fiber per gramm | 0.0000001 | $0.1*10^{-6}$ | 18.0 | 0.934 | 0.363 |
| age | -0.0000003 | $0.5*10^{-6}$ | 20.0 | -0.530 | 0.602 |
| sex (male) | 0.0000035 | $7.5*10^{-6}$ | 20.0 | 0.462 | 0.649 |
| BMI | -0.0000022 | $1.4*10^{-6}$ | 20.3 | -1.607 | 0.124 |
| time (follow-up) | -0.0000010 | $1.0*10^{-6}$ | 17.6 | -1.037 | 0.314 |
| intervention (fiber) | -0.0000001 | $1.0*10^{-6}$ | 17.6 | -0.115 | 0.910 |

#### **Extended Table 3-5d:** Coefficients of Model 7.2

| | Estimate | Std. Error | df | t value | Pr(>\|t\|) |
|---|---|---|---|---|---|
| intercept | 0.0010303 | $41.8*10^{-6}$ | 20.5 | 24.673 | 0.000 |
| fiber per 1000kcal | -0.0000003 | $0.3*10^{-6}$ | 17.8 | -1.097 | 0.287 |
| age | -0.0000002 | $0.5*10^{-6}$ | 19.9 | -0.450 | 0.657 |
| sex (male) | 0.0000035 | $7.5*10^{-6}$ | 19.9 | 0.463 | 0.648 |
| BMI | -0.0000026 | $1.4*10^{-6}$ | 20.2 | -1.877 | 0.075 |
| time (follow-up) | -0.0000011 | $1.0*10^{-6}$ | 17.5 | -1.094 | 0.289 |
| intervention (fiber) | -0.0000002 | $1.0*10^{-6}$ | 17.5 | -0.201 | 0.843 |

**Extended Table 3-5e:** Model comparison (ANOVA) of Model 7 and Model 7.1

| | npar | AIC | BIC | logLik | deviance | Chisq | Df | Pr(>Chisq) |
|---|---|---|---|---|---|---|---|---|
| fit7 | 8 | -3979 | -3952 | 1997 | -3995 | NA | NA | NA |
| fit7.1 | 9 | -3978 | -3948 | 1998 | -3996 | 0.883 | 1 | 0.348 |

**Extended Table 3-5f:** Model comparison (ANOVA) of Model 7 and Model 7.2

| | npar | AIC | BIC | logLik | deviance | Chisq | Df | Pr(>Chisq) |
|---|---|---|---|---|---|---|---|---|
| fit7 | 8 | -3979 | -3952 | 1997 | -3995 | NA | NA | NA |
| fit7.2 | 9 | -3978 | -3948 | 1998 | -3996 | 1.22 | 1 | 0.269 |

**Extended Table 6-1a:** Models for fiber intervention effects on TNF-α

| Model | Response | Fixed effects | Random effects |
|---|---|---|---|
| 2 | TNF-α | time+ intervention | 1 \| subject |
| 2.1 | TNF-α | time*intervention+ time+intervention | 1 \| subject |

**Extended Table 6-1b:** Coefficients of Model 2

| | Estimate | Std. Error | df | t value | Pr(>\|t\|) |
|---|---|---|---|---|---|
| intercept | 5.882 | 0.230 | 103 | 25.62 | 0.000 |
| time (follow-up) | 0.072 | 0.172 | 144 | 0.42 | 0.675 |
| intervention (fiber) | 0.081 | 0.173 | 145 | 0.47 | 0.639 |

**Extended Table 6-1c:** Coefficients of Model 2.1

| | Estimate | Std. Error | df | t value | Pr(>\|t\|) |
|---|---|---|---|---|---|
| intercept | 5.930 | 0.247 | 124 | 24.048 | 0.000 |
| time (follow-up) | -0.021 | 0.246 | 142 | -0.087 | 0.931 |
| intervention (fiber) | -0.008 | 0.240 | 147 | -0.034 | 0.973 |
| time (follow-up):intervention (fiber) | 0.185 | 0.345 | 143 | 0.536 | 0.593 |

**Extended Table 6-1d:** Model comparison (ANOVA) of Model 2 and Model 2.1

| | npar | AIC | BIC | logLik | deviance | Chisq | Df | Pr(>Chisq) |
|---|---|---|---|---|---|---|---|---|
| fit2 | 5 | 726 | 742 | -358 | 716 | NA | NA | NA |
| fit2.1 | 6 | 728 | 747 | -358 | 716 | 0.293 | 1 | 0.589 |

**Extended Table 6-2a:** Models for fiber intervention effects on CRP

| Model | Response | Fixed effects | Random effects |
|---|---|---|---|
| 4 | CRP | time+ intervention | 1 \| subject |
| 4.1 | CRP | time*intervention+ time+intervention | 1 \| subject |

**Extended Table 6-2b:** Coefficients of Model 4

| | Estimate | Std. Error | df | t value | Pr(>\|t\|) |
|---|---|---|---|---|---|
| intercept | 0.167 | 0.075 | 91.8 | 2.230 | 0.028 |
| time (follow-up) | -0.053 | 0.052 | 140.7 | -1.010 | 0.314 |
| intervention (fiber) | 0.003 | 0.052 | 141.7 | 0.056 | 0.955 |

**Extended Table 6-2c:** Coefficients of Model 4.1

| | Estimate | Std. Error | df | t value | Pr(>\|t\|) |
|---|---|---|---|---|---|
| intercept | 0.138 | 0.080 | 110 | 1.737 | 0.085 |
| time (follow-up) | 0.003 | 0.074 | 139 | 0.038 | 0.969 |
| intervention (fiber) | 0.056 | 0.073 | 144 | 0.770 | 0.442 |
| time (follow-up):intervention (fiber) | -0.110 | 0.104 | 140 | -1.051 | 0.295 |

**Extended Table 6-2d:** Model comparison (ANOVA) of Model 4 and Model 4.1

| | npar | AIC | BIC | logLik | deviance | Chisq | Df | Pr(>Chisq) |
|---|---|---|---|---|---|---|---|---|
| fit4 | 5 | 266 | 282 | -128 | 256 | NA | NA | NA |
| fit4.1 | 6 | 267 | 287 | -127 | 255 | 1.12 | 1 | 0.289 |

**Extended Table 6-3a:** Models for fiber intervention effects on hypothalamic MD

| Model | Response | Fixed effects | Random effects |
|---|---|---|---|
| 6 | MD_hyp | time+ intervention | 1 \| subject |
| 6.1 | MD_hyp | time*intervention+time+ intervention | 1 \| subject |

**Extended Table 6-3b:** Coefficients of Model 6

| | Estimate | Std. Error | df | t value | Pr(>\|t\|) |
|---|---|---|---|---|---|
| intercept | 0.0010036 | $5.6*10^{-6}$ | 30738451 | 180.794 | 0.000 |
| time (follow-up) | -0.0000013 | $1.8*10^{-6}$ | 30738451 | -0.681 | 0.496 |
| intervention (fiber) | -0.0000008 | $1.8*10^{-6}$ | 30738451 | -0.458 | 0.647 |

**Extended Table 6-3c:** Coefficients of Model 6.1

| | Estimate | Std. Error | df | t value | Pr(>\|t\|) |
|---|---|---|---|---|---|
| intercept | 0.0010020 | $5.6*10^{-6}$ | 1336524 | 177.648 | 0.000 |
| time (follow-up) | 0.0000019 | $2.6*10^{-6}$ | 1336524 | 0.741 | 0.459 |
| intervention (fiber) | 0.0000022 | $2.5*10^{-6}$ | 1336524 | 0.887 | 0.375 |
| time (follow-up):intervention (fiber) | -0.0000064 | $3.6*10^{-6}$ | 1336524 | -1.759 | 0.079 |

**Extended Table 6-3d:** Model comparison (ANOVA) of Model 6 and Model 6.1

| | npar | AIC | BIC | logLik | deviance | Chisq | Df | Pr(>Chisq) |
|---|---|---|---|---|---|---|---|---|
| fit6 | 5 | -3749 | -3732 | 1879 | -3759 | NA | NA | NA |
| fit6.1 | 6 | -3750 | -3730 | 1881 | -3762 | 3.12 | 1 | 0.077 |

**Extended Table 6-4a:** Hippocampal MD with intervention

| Model | Response | Fixed effects | Random effects |
|---|---|---|---|
| 8 | MD_hippo | time+ intervention | 1 \| subject |
| 8.1 | MD_hippo | time*intervention+time+intervention | 1 \| subject |

#### **Extended Table 6-4b:** Coefficients of Model 8

| | Estimate | Std. Error | df | t value | Pr(>\|t\|) |
|---|---|---|---|---|---|
| intercept | 0.0009522 | $3.3*10^{-6}$ | 21.7 | 289.153 | 0.000 |
| time (follow-up) | -0.0000010 | $1.0*10^{-6}$ | 18.2 | -0.976 | 0.342 |
| intervention (fiber) | 0.0000000 | $1.0*10^{-6}$ | 18.2 | -0.039 | 0.969 |

#### **Extended Table 6-4c:** Coefficients of Model 8.1

| | Estimate | Std. Error | df | t value | Pr(>\|t\|) |
|---|---|---|---|---|---|
| intercept | 0.0009523 | $3.3*10^{-6}$ | 22.0 | 285.891 | 0.000 |
| time (follow-up) | -0.0000011 | $1.4*10^{-6}$ | 18.2 | -0.805 | 0.431 |
| intervention (fiber) | -0.0000002 | $1.4*10^{-6}$ | 18.2 | -0.137 | 0.892 |
| time (follow-up):intervention (fiber) | 0.0000003 | $2.0*10^{-6}$ | 18.2 | 0.157 | 0.877 |

#### **Extended Table 6-4d:** Model comparison (ANOVA) of Model 8 and Model 8.1

| | npar | AIC | BIC | logLik | deviance | Chisq | Df | Pr(>Chisq) |
|---|---|---|---|---|---|---|---|---|
| fit8 | 5 | -3982 | -3965 | 1996 | -3992 | NA | NA | NA |
| fit8.1 | 6 | -3980 | -3960 | 1996 | -3992 | 0.025 | 1 | 0.874 |

#### **Extended Table 7-1a:** Effect of TNF-α on hypothalamic MD

| Model | Response | Fixed effects | Random effects |
|---|---|---|---|
| 11 | MD_hyp | age+sex+time+intervention + time*intervention | 1 \| subject |
| 11.1 | MD_hyp | TNF+ age+sex+time+intervention + time*intervention | 1 \| subject |

#### **Extended Table 7-1b:** Coefficients of Model 11

| | Estimate | Std. Error | df | t value | Pr(>\|t\|) |
|---|---|---|---|---|---|
| intercept | $988*10^{-6}$ | $25.4*10^{-6}$ | 1194693 | 38.920 | 0.000 |
| age | $0.60*10^{-6}$ | $0.90*10^{-6}$ | 1194693 | 0.721 | 0.471 |
| sex (male) | $-7.80*10^{-6}$ | $12.4*10^{-6}$ | 1194693 | -0.630 | 0.529 |

| | Estimate | Std. Error | df | t value | Pr(>\|t\|) |
|---|---|---|---|---|---|
| time (follow-up) | $2.30*10^{-6}$ | $2.60*10^{-6}$ | 1194693 | 0.883 | 0.377 |
| intervention (fiber) | $2.60*10^{-6}$ | $2.60*10^{-6}$ | 1194693 | 1.0171 | 0.309 |
| time (follow-up):intervention (fiber) | $-6.90*10^{-6}$ | $3.70*10^{-6}$ | 1194693 | -1.869 | 0.062 |

#### **Extended Table 7-1c:** Coefficients of Model 11.1

| | Estimate | Std. Error | df | t value | Pr(>\|t\|) |
|---|---|---|---|---|---|
| intercept | $994.8*10^{-6}$ | $25.6*10^{-6}$ | 240034.1 | 38.859 | 0.000 |
| TNF | $- 1.7*10^{-6}$ | $0.90*10^{-6}$ | 240034.1 | -1.899 | 0.058 |
| age | $0.7*10^{-6}$ | $0.90*10^{-6}$ | 240034.1 | 0.811 | 0.417 |
| sex (male) | $-6.8*10^{-6}$ | $12.4*10^{-6}$ | 240034.1 | -0.547 | 0.585 |
| time (follow-up) | $2.2*10^{-6}$ | $2.60*10^{-6}$ | 240034.1 | 0.860 | 0.390 |
| intervention (fiber) | $2.6*10^{-6}$ | $2.60*10^{-6}$ | 240034.1 | 1.028 | 0.304 |
| time (follow-up):intervention (fiber) | $-6.6*10^{-6}$ | $3.70*10^{-6}$ | 240034.1 | -1.795 | 0.073 |

#### **Extended Table 7-1d:** Model comparison (ANOVA) of Model 11 and Model 11.1

| | npar | AIC | BIC | logLik | deviance | Chisq | Df | Pr(>Chisq) |
|---|---|---|---|---|---|---|---|---|
| fit11 | 8 | -3651.794 | -3625.569 | 1833.897 | -3667.794 | NA | NA | NA |
| fit11.1 | 9 | -3653.468 | -3623.965 | 1835.734 | -3671.468 | 3.674 | 1 | 0.055 |

#### **Extended Table 7-2a:** Effect of CRP on hypothalamic MD

| Model | Response | Fixed effects | Random effects |
|---|---|---|---|
| 12 | MD_hyp | age+sex+time+intervention+ time*intervention | 1 \| subject |
| 12.1 | MD_hyp | CRP_log10+ age+sex+time+intervention+ time*intervention | 1 \| subject |

#### **Extended Table 7-2b:** Coefficients of Model 12

| | Estimate | Std. Error | df | t value | Pr(>\|t\|) |
|---|---|---|---|---|---|
| intercept | $988*10^{-6}$ | $25.4*10^{-6}$ | 1194693 | 38.920 | 0.000 |
| age | $60.0*10^{-6}$ | $0.90*10^{-6}$ | 1194693 | 0.721 | 0.471 |
| sex (male) | $-7.80*10^{-6}$ | $12.4*10^{-6}$ | 1194693 | -0.630 | 0.529 |
| time (follow-up) | $2.30*10^{-6}$ | $2.60*10^{-6}$ | 1194693 | 0.883 | 0.377 |
| intervention (fiber) | $2.60*10^{-6}$ | $2.60*10^{-6}$ | 1194693 | 1.017 | 0.309 |

| | Estimate | Std. Error | df | t value | Pr(>\|t\|) |
|---|---|---|---|---|---|
| time (follow-up):intervention (fiber) | $-6.90*10^{-6}$ | $3.70*10^{-6}$ | 1194693 | -1.869 | 0.062 |

#### **Extended Table 7-2c:** Coefficients of Model 12.1

| | Estimate | Std. Error | df | t value | Pr(>\|t\|) |
|---|---|---|---|---|---|
| intercept | $988.1*10^{-6}$ | $25.5*10^{-6}$ | 3657968 | 38.810 | 0.000 |
| CRP_log10 | $-0.3*10^{-6}$ | $2.90*10^{-6}$ | 3657968 | -0.101 | 0.919 |
| age | $0.6*10^{-6}$ | $0.90*10^{-6}$ | 3657968 | 0.721 | 0.471 |
| sex (male) | $-8.0*10^{-6}$ | $12.5*10^{-6}$ | 3657968 | -0.638 | 0.524 |
| time (follow-up) | $2.3*10^{-6}$ | $2.60*10^{-6}$ | 3657968 | 0.881 | 0.379 |
| intervention (fiber) | $2.6*10^{-6}$ | $2.60*10^{-6}$ | 3657968 | 1.018 | 0.309 |
| time (follow-up):intervention (fiber) | $-7.0*10^{-6}$ | $3.70*10^{-6}$ | 3657968 | -1.864 | 0.062 |

#### **Extended Table 7-2d:** Model comparison (ANOVA) of Model 12 and Model 12.1

| | npar | AIC | BIC | logLik | deviance | Chisq | Df | Pr(>Chisq) |
|---|---|---|---|---|---|---|---|---|
| fit12 | 8 | -3651.794 | -3625.569 | 1833.897 | -3667.794 | NA | NA | NA |
| fit12.1 | 9 | -3649.803 | -3620.300 | 1833.901 | -3667.803 | 0.009 | 1 | 0.925 |

#### **Extended Table 7-3a:** Habitual fiber intake and hypothalamic volume

| Model | Response | Fixed effects | Random effects |
|---|---|---|---|
| 9 | Hyp_vol | age+sex+BMI+eTIV+time+ intervention | 1 \| subject |
| 9.1 | Hyp_vol | fiber per gramm+age+sex+BMI+eTIV+ time+intervention | 1 \| subject |
| 9.2 | Hyp_vol | fiber per 1000kcal+age+sex+BMI+eTIV+ time+intervention | 1 \| subject |

#### **Extended Table 7-3b:** Coefficients of Model 9

| | Estimate | Std. Error | df | t value | Pr(>\|t\|) |
|---|---|---|---|---|---|
| intercept | 396.063 | 126.03 | 92.6 | 3.143 | 0.002 |
| age | 0.864 | 1.13 | 56.4 | 0.765 | 0.447 |
| sex (male) | 38.690 | 18.60 | 55.1 | 2.080 | 0.042 |
| BMI | 4.614 | 2.82 | 186.1 | 1.638 | 0.103 |

| | Estimate | Std. Error | df | t value | Pr(>\|t\|) |
|---|---|---|---|---|---|
| eTIV | 0.0001982 | 0.0000603 | 55.3 | 3.286 | 0.002 |
| time (follow-up) | 1.285 | 2.03 | 140.8 | 0.632 | 0.528 |
| intervention (fiber) | 2.673 | 2.03 | 141.0 | 1.315 | 0.191 |

**Extended Table 7-3c:** Coefficients of Model 9.1

| | Estimate | Std. Error | df | t value | Pr(>\|t\|) |
|---|---|---|---|---|---|
| intercept | 418.300 | 126.56 | 92.9 | 3.305 | 0.001 |
| fiber per gramm | -0.544 | 0.26 | 153.1 | -2.093 | 0.038 |
| age | 0.948 | 1.14 | 56.4 | 0.834 | 0.408 |
| sex (male) | 39.500 | 18.71 | 55.1 | 2.111 | 0.039 |
| BMI | 3.814 | 2.82 | 186.6 | 1.351 | 0.178 |
| eTIV | 0.0002017 | 0.0000607 | 55.2 | 3.323 | 0.002 |
| time (follow-up) | 1.238 | 2.00 | 139.8 | 0.618 | 0.538 |
| intervention (fiber) | 2.717 | 2.00 | 139.9 | 1.355 | 0.177 |

**Extended Table 7-3d:** Coefficients of Model 9.2

| | Estimate | Std. Error | df | t value | Pr(>\|t\|) |
|---|---|---|---|---|---|
| intercept | 410.956 | 126.725 | 93.6 | 3.24 | 0.002 |
| fiber per 1000kcal | -0.640 | 0.532 | 147.9 | -1.20 | 0.231 |
| age | 0.929 | 1.133 | 56.6 | 0.82 | 0.415 |
| sex (male) | 37.757 | 18.648 | 55.3 | 2.02 | 0.048 |
| BMI | 4.174 | 2.838 | 186.0 | 1.47 | 0.143 |
| eTIV | 0.0001997 | 0.0000605 | 55.3 | 3.30 | 0.002 |
| time (follow-up) | 1.178 | 2.031 | 139.9 | 0.58 | 0.563 |
| intervention (fiber) | 2.455 | 2.036 | 140.1 | 1.21 | 0.230 |

**Extended Table 7-3e:** Model comparison (ANOVA) of Model 9 and Model 9.1

| | npar | AIC | BIC | logLik | deviance | Chisq | Df | Pr(>Chisq) |
|---|---|---|---|---|---|---|---|---|
| fit9 | 9 | 1866 | 1895 | -924 | 1848 | NA | NA | NA |
| fit9.1 | 10 | 1863 | 1896 | -922 | 1843 | 4.37 | 1 | 0.036 |

**Extended Table 7-3f:** Model comparison (ANOVA) of Model 9 and Model 9.2

| | npar | AIC | BIC | logLik | deviance | Chisq | Df | Pr(>Chisq) |
|---|---|---|---|---|---|---|---|---|
| fit9 | 9 | 1866 | 1895 | -924 | 1848 | NA | NA | NA |
| fit9.2 | 10 | 1866 | 1899 | -923 | 1846 | 1.46 | 1 | 0.226 |

**Extended Table 7-4a:** Effect of supplementary fiber intake on hypothalamic volume

| Model | Response | Fixed effects | Random effects |
|---|---|---|---|
| 10 | Hyp_vol | time+ intervention | 1 \| subject |
| 10.1 | Hyp_vol | time*intervention+time+intervention | 1 \| subject |

**Extended Table 7-4b:** Coefficients Model 10

| | Estimate | Std. Error | df | t value | Pr(>\|t\|) |
|---|---|---|---|---|---|
| intercept | 886.35 | 8.82 | 60.8 | 100.484 | 0.000 |
| time (follow-up) | 1.23 | 2.04 | 140.5 | 0.603 | 0.548 |
| intervention (fiber) | 2.45 | 2.04 | 140.7 | 1.200 | 0.232 |

**Extended Table 7-4c:** Coefficients Model 10.1

| | Estimate | Std. Error | df | t value | Pr(>\|t\|) |
|---|---|---|---|---|---|
| intercept | 885.09 | 8.88 | 62.3 | 99.67 | 0.000 |
| time (follow-up) | 3.71 | 2.85 | 139.5 | 1.30 | 0.195 |
| intervention (fiber) | 4.89 | 2.83 | 140.2 | 1.73 | 0.086 |
| time (follow-up):intervention (fiber) | -5.08 | 4.08 | 139.6 | -1.25 | 0.215 |

**Extended Table 7-4d:** Model comparison (ANOVA) of Model 10 and Model 10.1

| | npar | AIC | BIC | logLik | deviance | Chisq | Df | Pr(>Chisq) |
|---|---|---|---|---|---|---|---|---|
| fit10 | 5 | 1890 | 1907 | -940 | 1880 | NA | NA | NA |
| fit10.1 | 6 | 1891 | 1911 | -939 | 1879 | 1.58 | 1 | 0.209 |

| | npar | AIC | BIC | logLik | deviance | Chisq | Df | Pr(>Chisq) |
|---|---|---|---|---|---|---|---|---|

### Computational Information

All calculations were performed using R programming language. For the detailed environment, please check the output of **sessionInfo()** below.

```
## R version 4.2.2 (2022-10-31)
## Platform: x86_64-pc-linux-gnu (64-bit)
## Running under: Debian GNU/Linux 11 (bullseye)
##
## Matrix products: default
## BLAS:
/afs/cbs.mpg.de/software/.r/4.2.2/debian-bullseye-amd64/lib/R/lib/
libRblas.so
## LAPACK:
/afs/cbs.mpg.de/software/.r/4.2.2/debian-bullseye-amd64/lib/R/lib/
libRlapack.so
##
## locale:
##  [1] LC_CTYPE=en_US.UTF-8       LC_NUMERIC=C
##  [3] LC_TIME=en_US.UTF-8        LC_COLLATE=en_US.UTF-8
##  [5] LC_MONETARY=en_US.UTF-8    LC_MESSAGES=en_US.UTF-8
##  [7] LC_PAPER=en_US.UTF-8       LC_NAME=C
##  [9] LC_ADDRESS=C               LC_TELEPHONE=C
## [11] LC_MEASUREMENT=en_US.UTF-8 LC_IDENTIFICATION=C
##
## attached base packages:
## [1] stats     graphics  grDevices utils     datasets  methods
base
##
## other attached packages:
##  [1] cowplot_1.1.1       jtools_2.2.1        gridExtra_2.3
##  [4] Rmisc_1.5.1         plyr_1.8.8          lattice_0.20-45
##  [7] lmerTest_3.1-3      performance_0.10.2  readxl_1.4.1
## [10] standardize_0.2.2   e1071_1.7-12        lm.beta_1.7-1
## [13] lme4_1.1-31         Matrix_1.5-1        writexl_1.4.2
## [16] moments_0.14.1      forcats_0.5.2       stringr_1.5.0
## [19] purrr_1.0.1         readr_2.1.3         tidyr_1.2.1
## [22] tibble_3.1.8        tidyverse_1.3.2     dplyr_1.0.10
## [25] magrittr_2.0.3      ggpubr_0.5.0        reshape2_1.4.4
## [28] openxlsx_4.2.5.1    ggplot2_3.4.0       psych_2.2.9
## [31] table1_1.4.3        broom.mixed_0.2.9.4
##
## loaded via a namespace (and not attached):
##  [1] googledrive_2.0.0   minqa_1.2.5         colorspace_2.0-3
##  [4] ggsignif_0.6.4      ellipsis_0.3.2      class_7.3-20
##  [7] fs_1.5.2            rstudioapi_0.14     proxy_0.4-27
## [10] farver_2.1.1        listenv_0.9.0       furrr_0.3.1
```

```
## [13] fansi_1.0.3          lubridate_1.9.0     xml2_1.3.3
## [16] codetools_0.2-18     splines_4.2.2       mnormt_2.1.1
## [19] knitr_1.41           Formula_1.2-4       jsonlite_1.8.4
## [22] nloptr_2.0.3         broom_1.0.2         dbplyr_2.3.0
## [25] compiler_4.2.2       httr_1.4.4          backports_1.4.1
## [28] assertthat_0.2.1     fastmap_1.1.0       gargle_1.2.1
## [31] cli_3.6.0            htmltools_0.5.4     tools_4.2.2
## [34] gtable_0.3.1         glue_1.6.2          Rcpp_1.0.9
## [37] carData_3.0-5        cellranger_1.1.0    vctrs_0.5.1
## [40] nlme_3.1-160         insight_0.18.8      xfun_0.36
## [43] globals_0.16.2       rvest_1.0.3         timechange_0.2.0
## [46] lifecycle_1.0.3      rstatix_0.7.1       googlesheets4_1.0.1
## [49] future_1.31.0        MASS_7.3-58.1       scales_1.2.1
## [52] hms_1.1.2            parallel_4.2.2      yaml_2.3.6
## [55] pander_0.6.5         stringi_1.7.12      highr_0.10
## [58] boot_1.3-28          zip_2.2.2           rlang_1.0.6
## [61] pkgconfig_2.0.3      evaluate_0.20       labeling_0.4.2
## [64] tidyselect_1.2.0     parallelly_1.34.0   R6_2.5.1
## [67] generics_0.1.3       DBI_1.1.3           mgcv_1.8-41
## [70] pillar_1.8.1         haven_2.5.1         withr_2.5.0
## [73] abind_1.4-5          modelr_0.1.10       crayon_1.5.2
## [76] car_3.1-1            utf8_1.2.2          tzdb_0.3.0
## [79] rmarkdown_2.20       grid_4.2.2          reprex_2.0.2
## [82] digest_0.6.31        xtable_1.8-4        numDeriv_2016.8-1.1
## [85] munsell_0.5.0
```
